# RANDOMIZED CONTROLLED TRIAL OF INTERMITTENT CALORIE RESTRICTION IN PEOPLE WITH MULTIPLE SCLEROSIS

**DOI:** 10.1101/2024.01.28.24301860

**Authors:** Laura Ghezzi, Valeria Tosti, Lisa Shi, Claudia Cantoni, Robert Mikesell, Samantha Lancia, Yanjiao Zhou, Kathleen Obert, Monokesh K. Sen, Anjie Ge, Miguel Tolentino, Bryan Bollman, Anthony S. Don, Giuseppe Matarese, Alessandra Colamatteo, Claudia La Rocca, Maria Teresa Lepore, Gregory F. Wu, Anne H. Cross, Robert T. Naismith, Luigi Fontana, Amber Salter, Laura Piccio

## Abstract

**Background:** Calorie restriction (CR) ameliorates preclinical models of multiple sclerosis (MS) through reduction of inflammation. The aim of this trial was to study the effects of 12-week iCR on metabolic, immunological and clinical outcomes in people with MS (pwMS).

**Methods:** Participants with relapsing-remitting MS were randomly assigned to intermittent CR (iCR) or a control group for 12 weeks. Primary outcome was change in leptin levels; secondary outcomes included changes in anthropometric and body composition measures, peripheral blood metabolic and immunologic profiling, and clinical measures. Mixed effects linear regression models were used to evaluate differences.

**Results:** Forty-two pwMS were randomized, 34 completed the study (17 iCR and 17 control). Leptin levels decreased in the iCR group and were significantly lower in the iCR than the control group at 6 (mean difference 11.49 μg/dL, 95% CI 32.54, 9.54; *P*=0.01) and 12 weeks (6.97 μg/dL, 95% CI 28.02, 14.06; *P*=0.03). We observed a significant reduction of weight, body mass index and body adiposity measures over the 6 and 12-weeks in the iCR group. Immune profiling showed a significant increase in CD45RO^+^ regulatory T cell numbers after 6 weeks of iCR. Lysophosphatidylcholine, lysophophatidylethanolamine and phosphatidylinositol lipid species were significantly increased after 12 weeks in the iCR group compared to baseline, and all three were higher at 12 weeks compared to controls. Exploratory cognitive testing demonstrated improvement in the symbol digit modality test score in the iCR group.

**Conclusions:** Short term iCR is safe, feasible and can benefit metabolic and immunologic profiles in pwMS.

ClinicalTrial.gov number: <u>NCT03539094</u> (first patient screened on 11/14/17; first patient recruited on 1/29/2018; last patient recruited on 11/24/2021).

## INTRODUCTION

Multiple sclerosis (MS) is an inflammatory, demyelinating, and neurodegenerative disease of the central nervous system (CNS). Despite treatments that control disease inflammatory activity, many still develop chronic progression, physical and cognitive disabilities, and significant personal and socio-economic burdens. The etiology of MS is unknown but involves genetic and environmental factors, among which diet is a possible contributor. Obesity early in life is associated with increased risk of developing MS ^1–3^, while obesity in people with MS (pwMS) correlates with worse progression and disability ^4^. Multiple mechanisms could underlie this association, including a role of adipose tissue in secreting pro-inflammatory adipokines, hormonal changes, and gut dysbiosis^5^. Calorie restriction (CR) is effective in ameliorating disability and dampening inflammation in MS preclinical models ^6^ ^7^. The mechanisms of CR are likely related to its anti-inflammatory and neuroprotective effects ^8^. However, research on CR in pwMS has been limited by practical difficulties. CR can be achieved by daily or intermittent reduction of calorie intake. Daily CR is difficult to implement and maintain in humans, while intermittent CR (iCR) may be more feasible.

Here, we conducted a randomized rater-blinded clinical study comparing the effects of iCR versus an unrestricted standard Western diet in people with relapsing-remitting MS (RRMS). The primary outcome of the study was change in serum levels of the adipokine leptin; secondary outcomes included changes in anthropometric and total body fat measures, peripheral blood metabolic and immunologic profiling.

## RESEARCH DESIGN AND METHODS

### Protocol Approvals, Registrations, and Patient Consents

The study was approved by the Washington University Institutional Review Board (#201707010). Written informed consent was signed by all participants. The study was registered at ClinicalTrials.gov (NCT03539094).

### Study subjects

pwMS were recruited at the John L. Trotter MS Center at Washington University School of Medicine (WUSM) beginning in January 2018 until November 2021. Enrollment criteria were age ≥ 18 years old, with confirmed RRMS based on 2010 McDonald criteria ^9^, Expanded Disability Status Scale (EDSS) <6.0 ^10^, and neurologically stable for 3 months preceding baseline visit. Patients were untreated or on a disease modifying therapy (DMT) for at least 3 months. Initially, only interferon-β or glatiramer acetate were allowed as DMT; subsequently, dimethyl fumarate, teriflunomide or natalizumab were also allowed. Participants were required to have body mass index (BMI) between 22 and 35 kg/m^2^ (eligibility widened to 22-38 kg/m^2^ during the study), stop any weight loss/special diet one month prior to baseline visit, not smoke tobacco or e-cigarettes, and have stable weight (<2 kg change) in the previous six months. Exclusion criteria included: other autoimmune diseases, chronic metabolic diseases (e.g. diabetes) or other physiological or pathological conditions (e.g. pregnancy or cancer) interfering with study assessments. Use of antibiotics was not allowed in the 3 months before the baseline visit. Use of systemic corticosteroids or omega-3/fish oil supplements were not allowed for 1 month before enrollment. Anticoagulant therapies (e.g. warfarin) or other drugs requiring monitoring of vitamin K intake were not allowed at any time. Use of other medications with no changes in dosage during the study was allowed.

### Study Design

A parallel, randomized, controlled clinical trial was conducted to examine the effect of iCR on serum leptin (Figure 1). MS participants were randomly allocated to one of two groups: iCR or control. The randomization allocation ratio was 1:1 and utilized a block randomization scheme, with investigators blinded to the block sizes. Complete outcome measurements were performed at baseline and week 12. At week 6 all assessments were performed, except oral glucose tolerance test (OGTT) and dual-energy X-ray absorptiometry (DEXA). Adverse events were monitored and reported.

**Figure 1.**
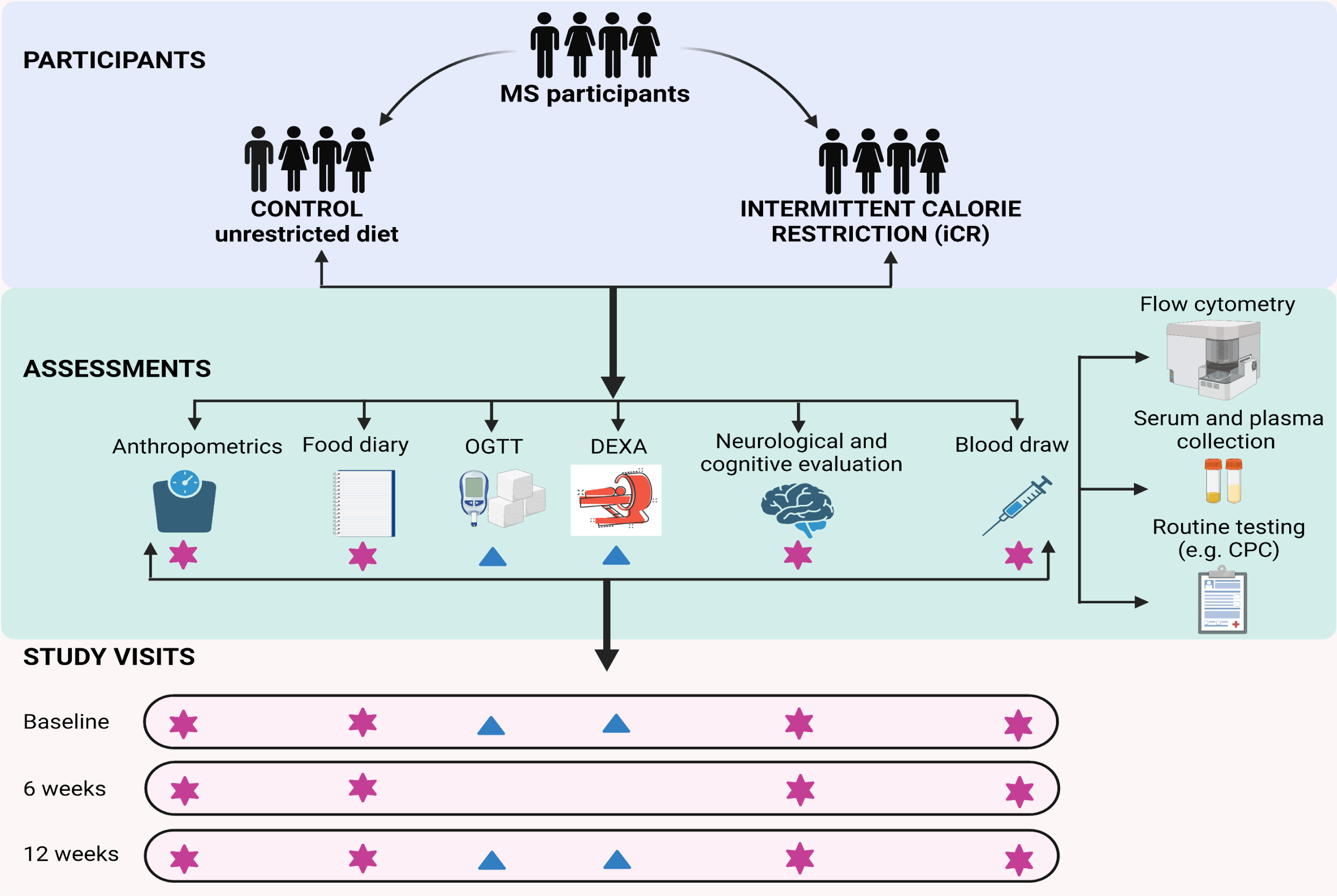
Overview of the study design.

### Intervention

Participants in the iCR group were instructed to restrict their calorie intake for two non-consecutive days per week. During fasting days, iCR group participants were requested to consume only non-starchy vegetables (raw or cooked), eaten plain or with oil (maximum 2 tablespoons/day), vinegar, lemon juice and/or seasonings, non-caloric drinks, not to exceed 500 calories/day. On non-fast days the iCR group was instructed to consume their usual food intake but to monitor portion sizes and food choices so as not to overeat. Participants randomized to the control group continued with their regular diet, with unrestricted access to food, but were asked to consume 1-1.5 cups of vegetables each day for the 12 weeks of the study in order to achieve similar consumption of vegetables in both groups. All study participants were asked to maintain their usual activity levels, which was monitored at baseline, 6-and 12-week visits using the physical activity recall (PAR) questionnaire administered by a trained team member using a standardized interview form ^11^.

### Study Outcomes

#### Adipokines and other analytes in blood

A venous blood sample was obtained after an overnight fast at baseline, 6, and 12 weeks, and aliquots of plasma and serum were frozen at -80°C. Samples were analyzed by the WUSM Core Laboratory for Clinical Studies in a single batch by technicians blinded to sample identity. The following analytes were measured in serum: leptin, high molecular weight (HMW) adiponectin, cortisol, insulin, IGF-1, β-hydroybutyrate, high sensitivity C-reactive protein (hsCRP), IL-6, neurofilament light chains (pNfL) (eMethods for details on the assays). Serum lipids, complete metabolic panel (CMP), cell blood count (CBC), and urinalysis were performed by the Core Laboratory on blood and urine collected at baseline, weeks 6 and 12.

#### Anthropometrics and body composition measures

Height, weight (measured using a Scale-Tronix portable scale, Welch Allyn, Inc.), and waist circumference were obtained with participants wearing only underwear and hospital gown, in the morning after 12h fast. A commercial scale (Tanita HD-662 digital scale) was provided for measuring body weight at home weekly. Body composition was evaluated by DEXA (Lunar iDXA, software version Encore 16v manufactured, GE Healthcare).

#### Oral glucose tolerance test (OGTT)

After an overnight fast and an adequate carbohydrate intake (at least 150 g/day) during the previous 3 days, subjects underwent a 2-hour OGTT with 75g oral glucose load (eMethods).

#### Dietary intake and adherence

Four-day food diaries (two weekdays and two weekend days) were collected to assess study eligibility at baseline and adherence to study interventions at week 6 and 12 in the iCR and control groups. One fasting day was included in the 4-day period for the iCR group to monitor their adherence to the fasting protocol. On a weekly basis, the study dietitian contacted participants (by phone or email) in both groups to collect morning weights, changes in medications, adverse events, and to provide motivational support for participants to follow their interventions. The weekly contact also served as a check on dietary adherence. Food diaries were analyzed to determine nutrient intakes using the NDSR program (Nutrition Data System for Research, University of Minnesota, version 2017 to version 2022 depending on the time when the food diary was collected).

The Healthy Eating Index (HEI) total score was calculated at each time point by NDSR as a measure of diet quality using 13 components to assess how well the diet aligns with recommendations of the 2015-2020 Dietary Guidelines for Americans ^12^ (eTable 1). A higher HEI total score indicates a diet that aligns better with the recommendations. In the iCR group, adherence to the intervention was calculated as a percentage with the number of fast days completed divided by the total number of expected fast days during the study; this fraction was then multiplied by 100. The degree of CR was calculated by subtracting the calorie intake at week 6 or week 12 from calorie intake at baseline, divided by the calorie intake at baseline; this fraction was multiplied by 100.

#### Blood immune cell subsets

Flow cytometric analysis was performed on fresh heparinized whole blood as previously described ^6^. Samples were run on a Gallios flow cytometer (Beckman Coulter); analysis was done with Kaluza Software (version 2.0, Beckman Coulter). Antibodies and gating strategy are shown in eTable 2a and Suppl. Figure 1, respectively.

#### Western blot analysis

Total cell lysates of regulatory T cells (Tregs) and effector T cells (Teffs) were obtained after 20 min incubation at 4°C in RIPA buffer (Sigma-Aldrich), plus SIGMAFAST Protease Inhibitor and Sigma Phosphatase Inhibitor (Sigma-Aldrich). Western blot analysis was performed as described ^13^ ^14^. Membranes were quantified by densitometric analysis using ImageJ 1.47v. Densitometric results are reported as fold over baseline.

#### Lipidomics

Lipids were extracted from 20 µL plasma samples collected at baseline and week 12 using the two-phase methyl-*tert*-butyl ether (MTBE) method, with inclusion of an internal standard for each lipid class ^15^. Lipids were quantified by liquid chromatography-tandem mass spectrometry (LC-MS/MS), using selected reaction monitoring on a TSQ Altis mass spectrometer with Vanquish HPLC. HPLC conditions were as described ^16^. Precursor and product ion pairs are listed in eTable 2c. Peaks were integrated using Tracefinder 5.1 software and lipids were quantified by normalisation to class-specific internal standards.

#### Neurological assessments and patient reported outcomes (PROs)

The EDSS ^10^ was performed at baseline, 6 and 12 week visit by a neurologist blinded to the intervention. Standardized tests of manual dexterity (9-hole peg test 9HPT, each hand), timed 25 foot-walk and cognitive functions (Symbol Digit modality test, SDMT) ^17^ were performed along with PROs including the MS impact scale-29 (MSIS) physical and mental subscales ^18^ and the modified fatigue impact scale (MFIS) psychosocial, cognitive and physical subscales ^19^.

### Statistical Analyses

Descriptive statistics were used to summarize demographic and clinical characteristics and differences between groups evaluated using a t-test, Mann Whitney U test or chi square test, as appropriate. The intention to treat (ITT) principle was used for all analyses. We evaluated differences in the outcomes of interest using mixed effects linear regression models. The model included three time points (baseline, 6 weeks and 12 weeks) evaluating between group differences (iCR vs Control) and changes within group over time. An interaction term was used to evaluate overall changes between groups and each time point. Additionally, the models were adjusted for age, sex, and DMT use. Model assumptions were verified, and violations were addressed using transformations. Laboratory data were log transformed for analysis due to the data violating the normality assumption. Raw means and differences are reported; the *P* values are based on the multivariable model adjusting for age, sex and DMT use. The significance level for the primary outcome was 0.05. A 5% false discovery rate (FDR) correction was implemented to control for multiple comparison among secondary and exploratory outcomes. The significance level associated with the FDR correction was p<0.0013. Data analyses were conducted in SAS v9.4.

## RESULTS

### Characteristics of study subjects

Sixty-one pwMS were consented (n=8 men; n=53 women) and 42 participants were randomized; 22 were assigned to the iCR and 20 to the control group. Baseline demographic, anthropometric, and clinical characteristics of the two groups were similar (Table 1). Five participants withdrew from the iCR group, while 3 withdrew from the control group during the intervention. Thirty-four of the 42 enrolled subjects (80.9%; 17 participants/group) completed the study (Figure 2; CONSORT diagram).

**Table 1:** Baseline Characteristics of MS participants

| Factor | Total<br>(N=42) | iCR<br>(N=22) | Control<br>(N=20) | p-value |
| --- | --- | --- | --- | --- |
| **Race, No. (%)** |  |  |  | 0.35<sup>F</sup> |
| · Caucasian | 36(85.7) | 19(86.4) | 17(85.0) |  |
| · African American | 4(9.5) | 1(4.5) | 3(15.0) |  |
| · Other | 2(4.8) | 2(9.1) | 0(0.0) |  |
| **Ethnicity, No. (%)** |  |  |  | 0.48<sup>F</sup> |
| · Not Hispanic | 41(97.6) | 22(100.0) | 19(95.0) |  |
| · Hispanic or Latino | 1(2.4) | 0(0.0) | 1(5.0) |  |
| **Gender/Sex, No. (%)** |  |  |  | 0.40<sup>F</sup> |
| · Male | 6(14.3) | 2(9.1) | 4(20.0) |  |
| · Female | 36(85.7) | 20(90.9) | 16(80.0) |  |
| **Age at Consent (years), Mean ± SD** | 48.2±9.8 | 50.0±9.5 | 46.4±10.1 | 0.24 |
| **Disease Duration (years), Median (Q1, Q3)** | 11.0[5.0,19.0] | 15.0[8.0,21.0] | 7.5[4.5,14.0] |  |
| **Number of relapses\*, Median (Q1, Q3)** | 1.00[0.00,2.0] | 2.0[0.00,5.0] | 1.00[0.00,2.0] |  |
| **EDSS, Median (Q1, Q3)** | 2.0[1.5,2.5] | 2.0[1.5,2.5] | 2.0[1.3,2.5] |  |
| **Body Mass Index, Mean ± SD** | 28.7±4.3 | 27.9±3.6 | 29.6±4.9 | 0.20 |
| **Weight in kg, Mean ± SD** | 80.7±15.2 | 78.3±13.1 | 83.4±17.1 | 0.28 |
| **Waist circumference in centimeters, Mean ± SD** | 99.3±11.4 | 97.5±10.7 | 101.4±12.0 | 0.27 |
| **DMT, No. (%)** |  |  |  | 0.15<sup>F</sup> |
| · None | 3(7.1) | 3(13.6) | 0(0.0) |  |
| · Avonex | 5(11.9) | 2(9.1) | 3(15.0) |  |
| · Copaxone | 15(35.7) | 10(45.5) | 5(25.0) |  |
| · Rebif | 6(14.3) | 1(4.5) | 5(25.0) |  |
| · Tecfidera | 6(14.3) | 4(18.2) | 2(10.0) |  |
| · Plegridy | 2(4.8) | 0(0.0) | 2(10.0) |  |
| · Tysabri | 3(7.1) | 1(4.5) | 2(10.0) |  |
| · Aubagio | 2(4.8) | 1(4.5) | 1(5.0) |  |
| **DMT Class, No. (%)** |  |  |  | 0.87 |
| · None/Injectable | 31(73.8) | 16(72.7) | 15(75.0) |  |
| · Oral/Tysabri | 11(26.2) | 6(27.3) | 5(25.0) |  |

\*Data not available for all subjects. Missing values: Number of relapses = 4.
Values presented as Mean ± SD with ANOVA, Median [P25, P75] or Median (min, max) with Kruskal-Wallis test, or N (column %) with Fisher's Exact test (F) or Pearson's chi-square test.

**Figure 2.**
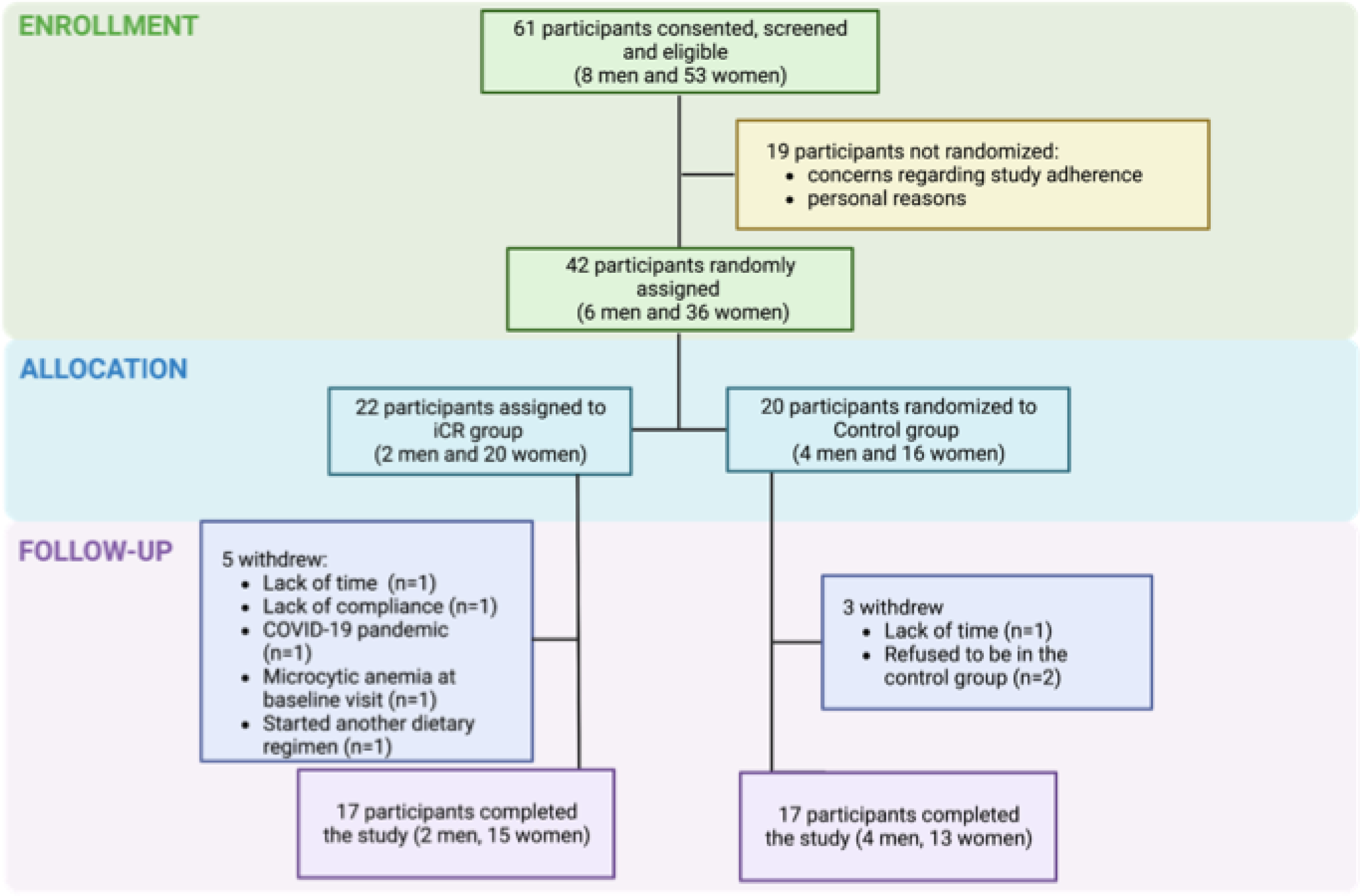
CONSORT Flow Diagram.

### Adherence to the iCR regimen and dietary intake

Adherence to the two fast days per week was 99.5% for the first 6 weeks and 97.2% for the second 6 weeks of the study. In the iCR group, calorie intake at week 6 and week 12 resulted in an overall calorie restriction of 21.5% and 23.1%, compared to baseline (eTable 4), confirming adherence to expected CR.

### Adverse events and safety of iCR in relapsing MS

No adverse events grade 3 or higher (using Common Terminology Criteria for Adverse Events-version 5.0) were seen in either group. Three of the 22 iCR group participants (13.6%) reported mild symptoms determined to be probably related to the intervention, including headache, lightheadedness, tiredness, bloating and loose stool on fasting days. No enrolled subjects changed DMT during the study. One control group participant developed optic neuritis, treated with corticosteroids. CBC, CMP and urinalysis performed at all visits did not reveal any negative effects of iCR.

### Reduction of leptin levels at 6 and 12 weeks and increased adiponectin with iCR

In the iCR group, leptin levels decreased at 6 weeks (mean leptin difference 11.49 μg/dL, 95%CI 32.54, 9.54; *P*=0.01) and 12 weeks (6.97 μg/dL, 95%CI 28.02, 14.06; *P*=0.03) compared to the control group. Compared to baseline, mean reduction of leptin in the iCR group was 2.85 μg/dL (95% CI -4.18, 9.89; *P*=0.04) at 6 weeks, and 2.2 μg/dL (95% CI -4.83, 9.24; *P*=0.06) at 12 weeks (Table 2). Leptin levels in the control group did not change at 6 and 12 weeks from baseline (Table 2; Figure 3A). In the iCR group, high molecular weight adiponectin increased 536 ng/ml (95% CI 1042, 30; *P*=0.02) at 6 weeks, and 610 ng/ml (95% CI 1116, 104; *P*=0.01) at 12 weeks compared to baseline, while no significant changes were observed over time in the control group (Table 2; Figure 3A).

**Table 2:**
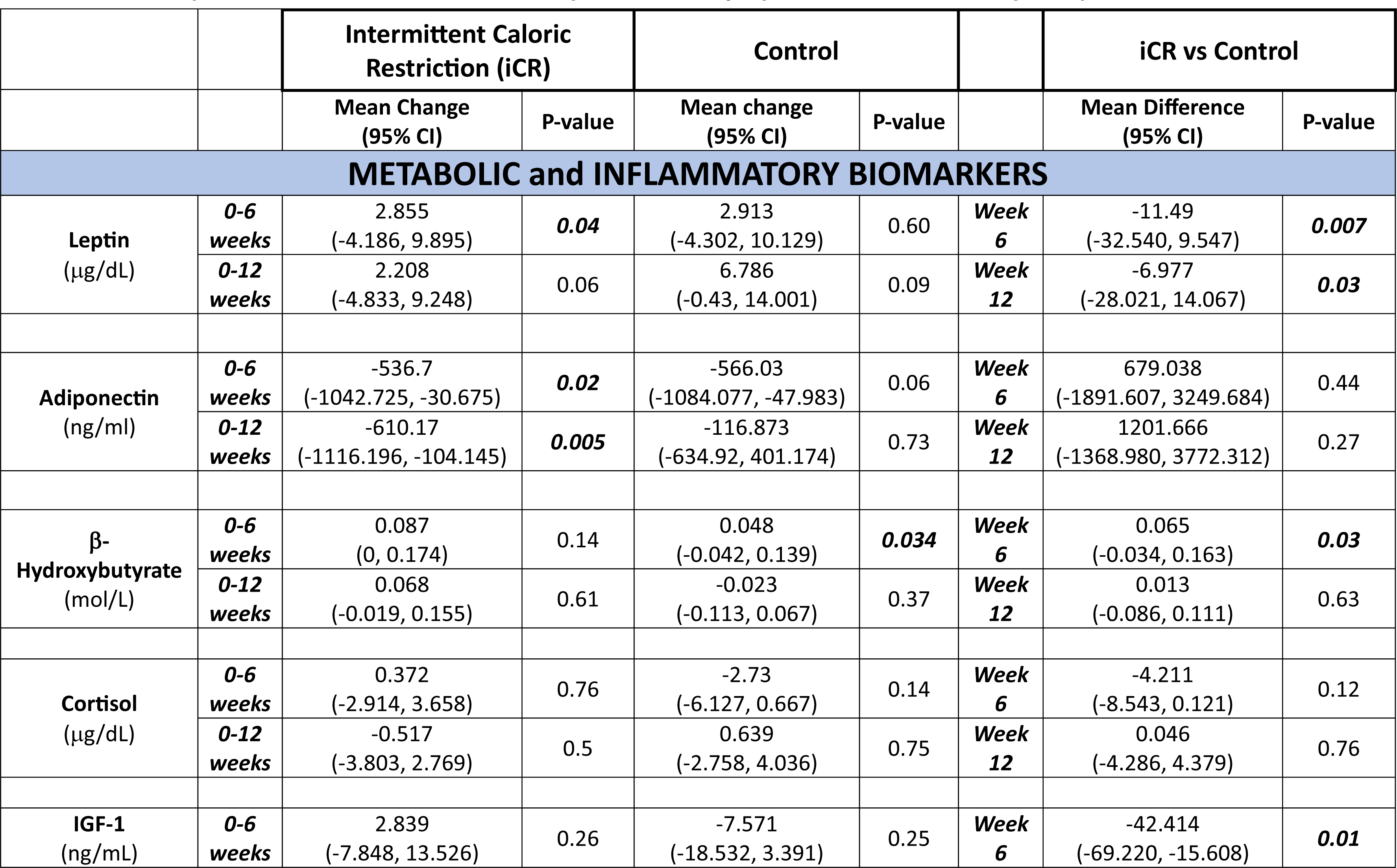
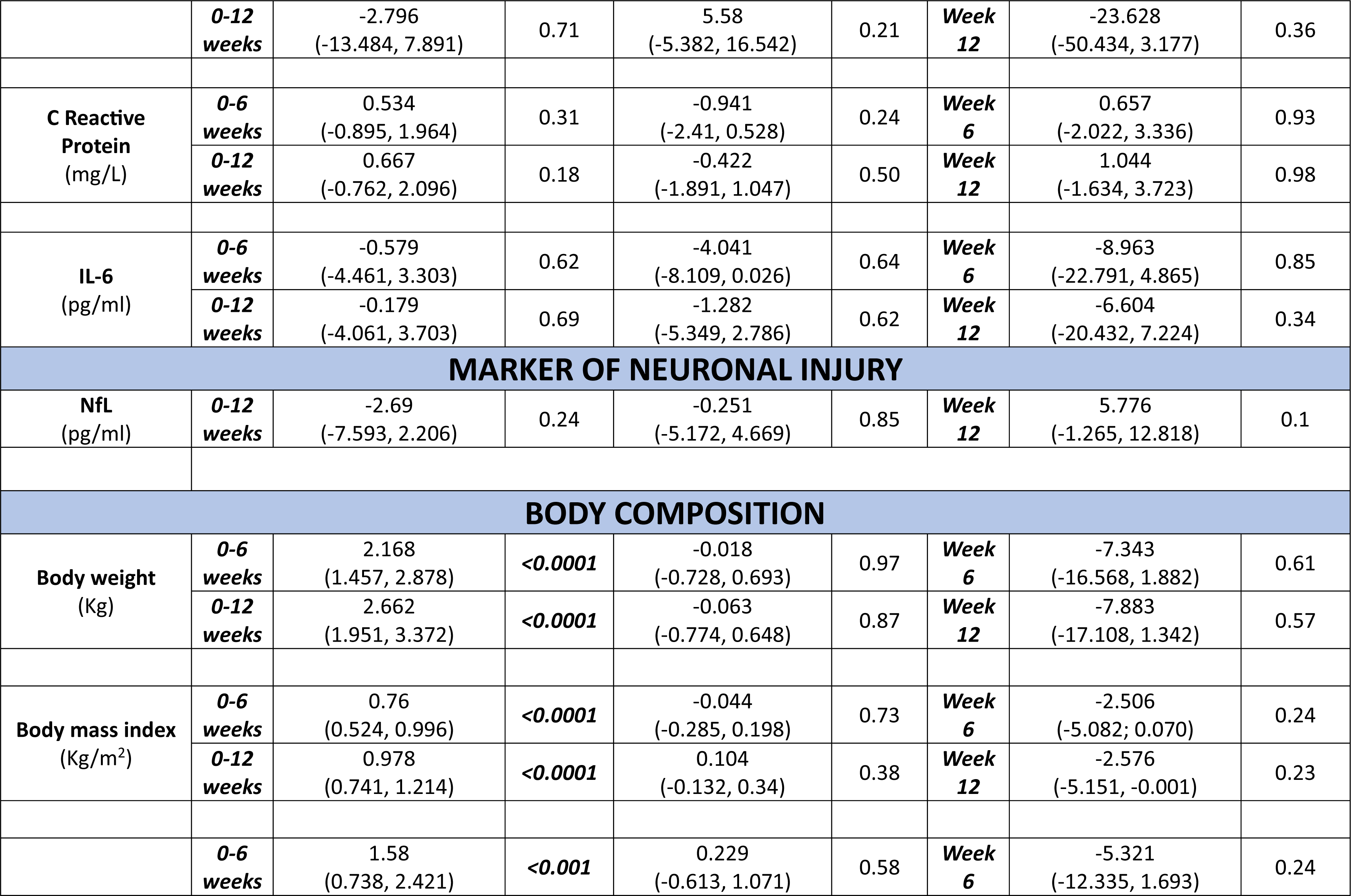
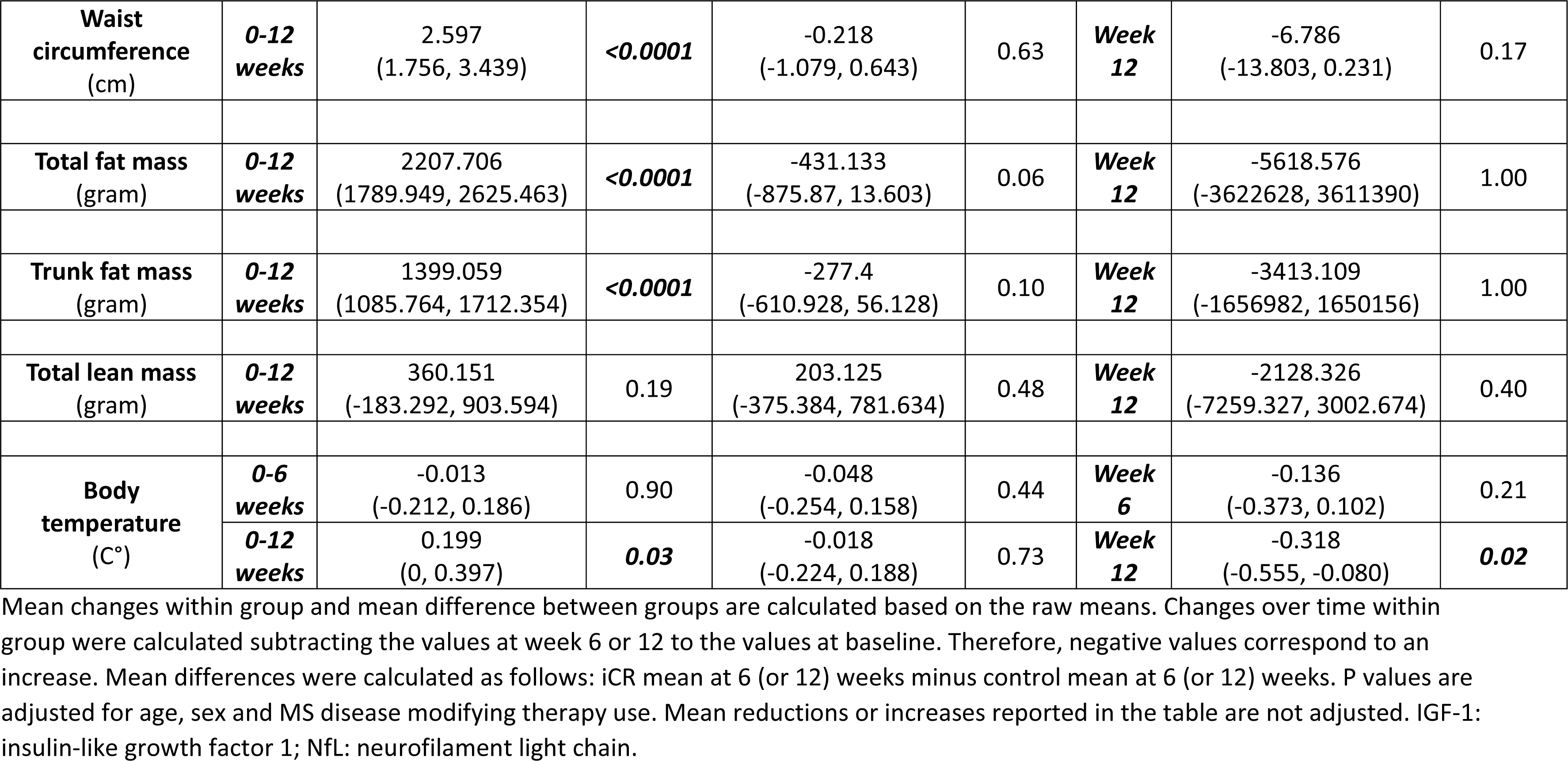
Adipokines, metabolic, inflammatory, neuronal injury biomarkers and body composition measures.

**Figure 3.**
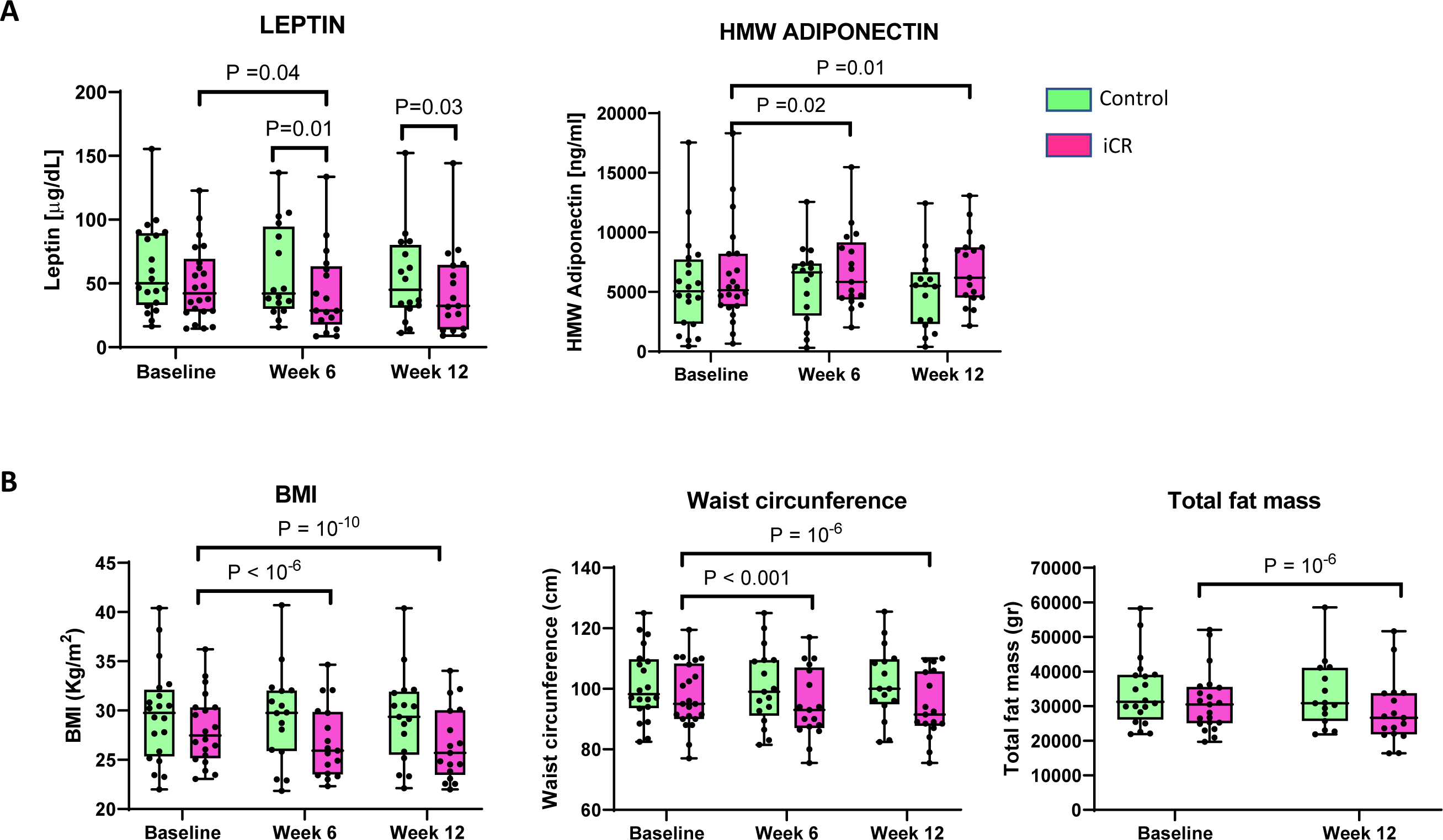
Changes in adipokine and anthropometric measures in the iCR and control groups over the course of the study. (A) Leptin and HMW adiponectin serum levels at 6 and 12 weeks. (B) BMI, waist circumference and total fat measured by DEXA at 6 and 12 weeks. Individual values for each subject are reported in the graph. The box extends from the 25th to 75th percentiles. Horizontal bars are median and error bars are min and max values; HMW=high molecular weight; BMI=Body Mass Index.

Differences in serum levels of adiponectin did not remain statistically significant at the FDR adjusted significance level. The iCR group had higher levels of βhydroxybutyrate compared to control which reached statistical significance at 6 weeks (*P*=0.03). Levels of IGF-1 were significantly lower at 6 weeks in the iCR group (*P*=0.01). No changes in cortisol, IL-6, CRP and pNfL were observed over time and between the two groups (Table 2).

### Reduction of anthropometric and body adiposity measures after 12-weeks of iCR

The iCR group participants showed a continual and significant decrease in weight, BMI, and waist circumference throughout the study, after adjusting for age, sex and DMT (Table 2). The iCR group lost a mean of 2.16 Kg (95%CI: 1.45-2.87; P<0.0001) at week 6 and 2.66 Kg (95%CI: 1.95-3.37; *P*<0.0001) at week 12 compared to baseline, with a mean decrease in BMI of 0.76 kg/m^2^ (95%CI 0.52-0.99; *P*<0.0001) and 0.97 kg/m^2^ (95%CI: 0.74-1.21; *P*<0.0001) at week 6 and 12, respectively. These results remained statistically significant at the FDR-adjusted significance level. No significant changes in weight, BMI, or waist circumference were observed in the control group over time (Table 2; Figure 3B). No significant differences between iCR and control groups at week 6 or week 12 were detected for weight, BMI, and waist circumference.

Body composition studies were performed by DEXA at baseline and week 12. Total and trunk fat were significantly reduced at week 12 compared to baseline in the iCR group with a mean decrease of 2.2 Kg in total body fat (95%CI: 1.78-2.62; *P*<0.0001) and 1.39 Kg in trunk fat (95%CI: 1.08-1.71; P<0.0001). These results remain statistically significant at the FDR adjusted significance level. No changes over time in the control group for body composition measures were observed. Additionally, no differences were noted between the two diet groups at weeks 6 and 12. Total lean mass and bone mass did not change over time within each group or between groups at 12 weeks (Table 2).

### Increased HDL levels after 6 and 12 weeks of iCR

HDL levels increased significantly in the iCR group at weeks 6 and 12 (mean increase of 5.91 mg/dL, 95%CI: -8.52-3.32; *P*<0.0001 and 2.32, 95%CI: -4.93, 0.28; *P*=0.04, respectively), while the control group exhibited no change over time (eTable 4). No differences in HDL levels were observed between the two groups at week 6 or 12.

Triglycerides, total cholesterol, and LDL levels did not significantly differ between the iCR and control groups at 6 and 12 weeks. These did not change over time within either group, apart from a reduction in triglycerides levels in the iCR group at week 6 (mean reduction 23.9 mg/dL, 95%CI 6.4-41.4; P=0.01). No clinically significant effects by iCR on glucose metabolism were observed. None of these differences remained significant after FDR adjustment.

### Twelve weeks of iCR altered circulating T cell subsets

Flow cytometric phenotyping of peripheral blood immune cells was performed at baseline and weeks 6 and 12 (eTable 5). While no significant changes were observed in total CD4+ and CD8+ T lymphocytes, some T cell subsets displayed intergroup differences (Figure 4). At 12 weeks higher numbers of naïve CD4+ T cells were observed in the iCR compared to the control group (iCR mean 241.8 cell /uL, 95%CI: 176.6, 07.1; control mean 112, 95%CI: 44.2, 179.9 *P*=0.04). Similar changes were observed for percentages of naïve CD4+ T cells (Figure 4; eTable 5).

**FIGURE 4.**
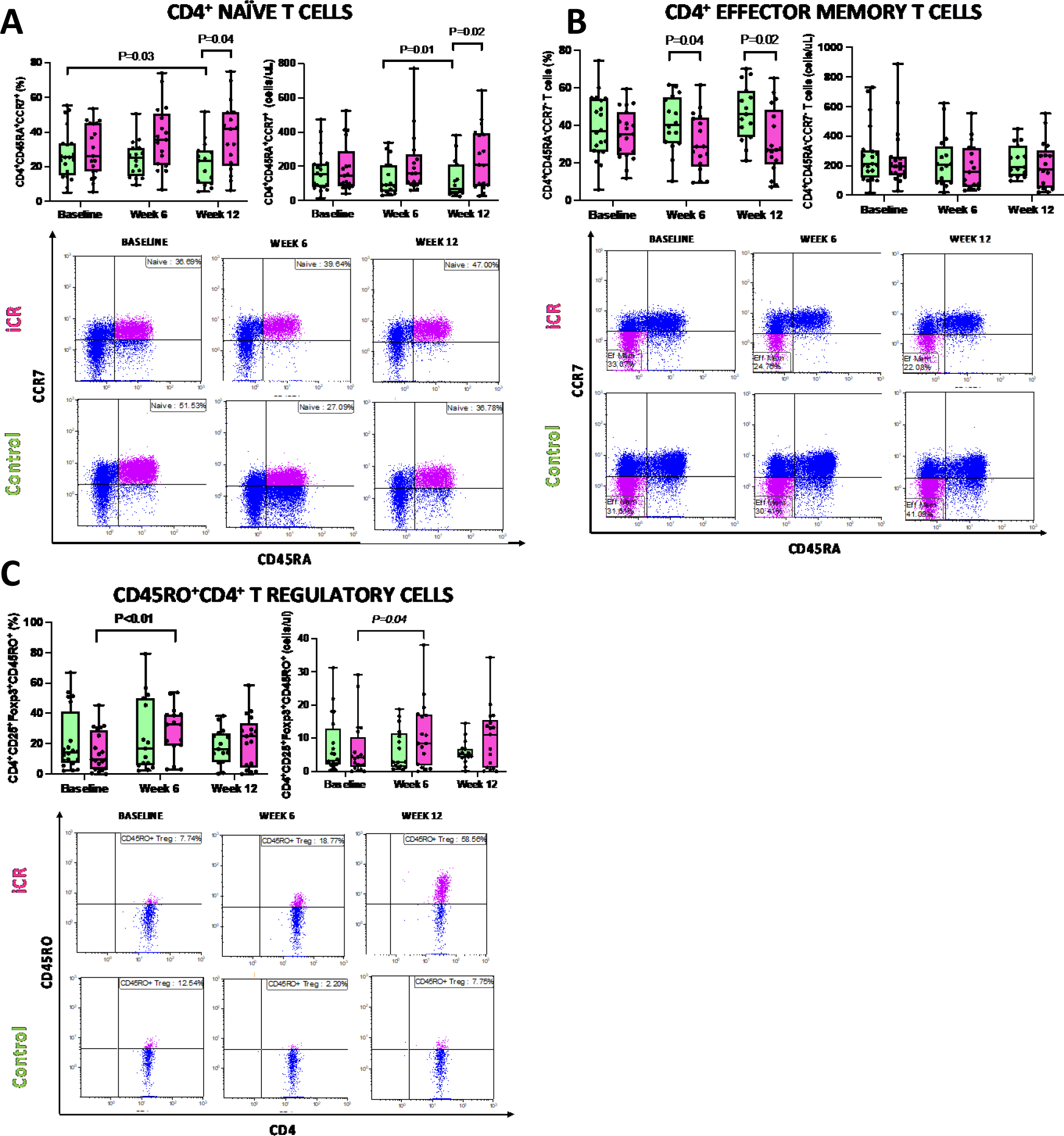
Changes in T cell subsets in the iCR and control groups over the course of the study. (A) Absolute numbers and frequencies of (A) naïve CD4+ T cells, (B) effector memory CD4+ T cells and (C) CD45RO+ T regulatory cells at 6 and 12 weeks in the iCR and control groups. Individual values for each subject are reported in the graph. The box extends from the 25th to 75th percentiles. Horizontal bars are median and error bars are min and max values. Representitive flow cytometry plots from the iCR and control groups are reported for each immune cell subset at each time point (the pink colored dots identify the subset quantified in the corresponding graphs).

Effector memory CD4+ T cell percentages decreased at the 6 and 12 week time points in the iCR compared to the control group (*P*=0.04 and *P*=0.02, respectively). Th1 cell numbers decreased in the iCR group over the study course (mean decrease at 6 weeks: 66.8 cell/μL, 95%CI: 15.3, 118.1; P=0.02; change from baseline to 12 weeks: 53.5, 95%CI: 2.1, 104.9; *P*=0.07) and were significant lower in the iCR compared to the control group at week 6 (iCR mean: 67.3 cell/μL, 95%CI: 24.1, 110.6; control mean: 80.6, 95%CI: 36.1, 125.2; *P*=0.02) (eTable 5). CD45RO+ T regs increased in the iCR group between baseline and 6- and 12-weeks, which reached statistically significance at 6 weeks for both absolute values and percentages (mean increase at 6 weeks: -4.9 cells/uL, 95%CI: -9.7, -0.2, *P*=0.04; -14.1 %, 95%CI: -25.7, -2.5, *P*=0.03), while no differences were noted between the iCR and control groups at either time point.

Since Teffs and Tregs engage different metabolic pathways, including glycolysis, lipid synthesis and fatty acid oxidation (FAO), to sustain their own proliferation and function ^20–22^, we evaluated levels of several enzymes that regulate these metabolic pathways. Tregs isolated from iCR group showed a significant increase of the glycolytic enzyme, Enolase-1, compared to control group; a similar trend was observed for Hexokinase II and Aldolase at 6 and 12 weeks (eFigure 2). Moreover, for FAO and lipid synthesis, we observed a significant increase of ACAD9 and FAS levels in Tregs from iCR compared to the control group, at 6 and 12 weeks, respectively; CPT1a and Apo-A4 showed a similar trend, in particular at 6 weeks (eFigure 2). On the contrary, we didn’t observe a clear change in specific metabolic pathways engaged by Teff cells (eFigure 3). However, Teff cells from the iCR group appeared less metabolically active compared to the control group.

At 12 weeks percentages of mucosal associated T cells (MAIT) cells were significantly higher in the iCR compared to the control group (iCR mean 13.6 %, IC: 9.5, 17.8; control mean: 11 %; IC: 6.8, 15.3; *P*=0.02) . Naive B cell numbers also trended to be higher in the iCR group compared to controls (iCR mean 66.6 cells/ml, 95% IC: 37.2, 95.978, P=0.06; control mean: 35.7, 95% IC: 4.5, 66.9, *P*=0.05) (eTable 5). No significant effects of iCR were detected on the other lymphoid and myeloid cell subsets analyzed.

### Twelve weeks of iCR altered circulating lipids

A total of 159 phospholipid, sphingolipid, acylcarnitine and diacylglycerol (DG) species were quantified in baseline and 12-week samples using LC-MS/MS (eTable 6). After FDR correction, nine lipids were significantly increased after 12 weeks of iCR, whereas no lipids changed in the control group (FDR-corrected P<0.05) (Figure 5 A, B; eTable 7). Specifically, five lysophosphatidylcholine [LPC(16:0), (18:1), (18:2), (20:4) and (22:4)], two lysophophatidylethanolamine [LPE(16:0) and (18:1)] and two phosphatidylinositol [PI(18:1/18:2) and (18:1/20:4)] species were significantly increased in the iCR cohort. Three of these [LPC(22:4), LPE(16:0) and PI(18:1/20:4)] were also higher in the iCR vs. control groups at 12 weeks (Figure 5C, E-G), as well as PI(16:0/18:1), PI(16:0/20:4), PI(16:1/18:1), LPE(18:0) and DG(18:0/18:1) (FDR-corrected P<0.05). Two sphingolipids, hexosylceramide (HexCer(d18:1/22:1)) and lactosylceramide (LacCer(d18:1/22:1)), were decreased after 12 weeks of iCR compared to the control group (FDR-corrected P<0.05) (Figure 5C). No lipids differed significantly between the groups at baseline (Figure 5D). Correlation analyses between the 16 lipids altered by iCR and other outcomes significantly affected by the diet at 12 weeks (Figure 5H; Suppl. Table 8) revealed MFIS to be associated with LPE (18:1) after correcting for FDR (psychosocial subscale; *r*=0.6, FDR-corrected *P=0.03*).

**FIGURE 5.**
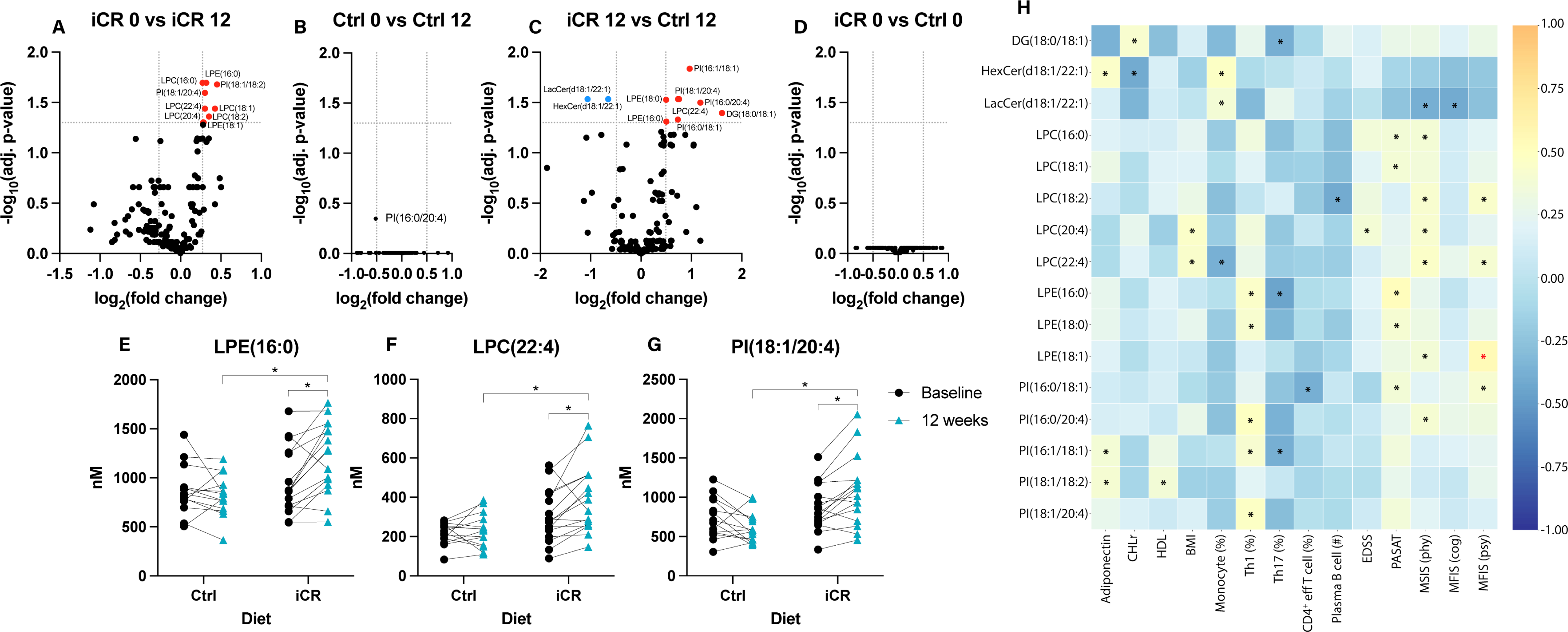
LPC, LPE, and PI species are affected by iCR. (A,B) Volcano plots showing effect of (A) iCR or (B) control diet on lipid levels at 12 weeks compared to baseline. (C,D) Comparison of lipid levels between iCR and control groups at (C) 12 weeks and (D) baseline. P values are adjusted for FDR. (E-G) Significant change in levels of (E) LPE(16:0), (F) LPC(22:4), and (G) PI(18:1/20:4) between baseline (closed circle) and 12 weeks (triangle) and within diet groups (* indicates significance after FDR adjustment). (H) Correlation heatmap illustrating significant correlations between the 16 lipids that were significantly altered after iCR and laboratory/clinical variables that were significantly altered after iCR. * = *P*<0.05, * indicates significance after FDR adjustment. DG, diacylglycerol; HexCer, hexosylceramide; LacCer, lactosylceramide; LPC, lysophosphatidylcholine; LPE, lysophosphatidylethanolamine; PI, phosphatidylinositol; CHDLr, cholesterol to high-density lipoprotein ratio; eff: effector.

### Measures of cognitive function and fatigue improved after twelve weeks of iCR

Standardized clinical testing assessed neurological disability, cognitive performance, and PROs at baseline, 6 and 12 weeks (eTable 9). These outcomes were exploratory given the short study duration, subjective nature of some of the measures, and the fact that patients were unblinded to the intervention. The SDMT score increased in the iCR group at the 6 (mean increase of 3.5, 95%CI: 0.6, 6.3; P=0.01) and 12 week time points (mean increase of 6.2, 95%CI 3.4, 9.5; P<0.0001) compared to baseline and was higher in the iCR versus control group at 12 weeks (*P*=0.03) (eFigure 4; eTable 9). No significant differences were observed in the control group over time. The MFIS scores (a PRO) decreased over the 12-week intervention in the iCR group for both the cognitive and psychosocial subscales (respective reductions of mean 2.5, 95%CI 0.3, 4.7, P=0.02; and 2.8, 95%CI 0.8, 4.8; *P*=0.006). No differences were observed in the control group. The iCR group MSIS score improved from baseline to week 6 and 12 in the mental subscale (mean change 0-12 weeks: 7.36, 95%CI: 3.61, 11.1*, P*=0.0002) and from baseline to week 6 in the physical scale (mean change: 2.7, 95%CI: 0.41, 5.12*, P*=0.02), with no changes over time in the control group. We did not observe any clinically significant changes in the EDSS, 9HPT, PASAT or T25W scores (eTable 9). Clinical differences were no longer statistically significant after FDR adjustment.

## DISCUSSION

PwMS are interested in potential lifestyle modifications, including diet changes ^23^, yet rigorous scientific evidence to guide clinicians and patients is limited. The current randomized controlled clinical trial in pwRRMS had the overall goal to address this unmet need by assessing the effects of iCR on metabolic, immunologic, and clinical measures compared to a control group eating an unrestricted diet. MS is associated with immune cell activation and neuroinflammation; RRMS subjects undergoing 12-weeks of iCR had reduced leptin levels, an adipokine with a known role in activating immune cells and inflammation. This trial and several prior clinical studies in pwMS indicate that daily or intermittent CR is feasible and improves subjective clinical measures ^6^ ^24^ ^25^. Furthermore, it adds to existing literature by showing objective effects on metabolic and immunologic biomarkers, not previously extensively investigated and with a potential beneficial impact on disease pathogenic mechanisms.

This study met its pre-specified primary outcome of a reduction of leptin levels with iCR. Leptin is an adipokine released by the adipose tissue proportionally to body fat, and which has pro-inflammatory effects. ^26^. Herein, reduction of leptin and increase in adiponectin occurred concomitantly to significant decreases in body weight, BMI, and body fat measures after 6 and 12-weeks of iCR. No major effects on markers of systemic inflammation (e.g.CRP and IL-6) were observed. In contrast to our trial, an 8-week study in pwMS comparing daily CR and iCR (two fasting days/week) with *ad libitum* feeding (n=12/group) did not report significant changes in circulating leptin and adiponectin, despite weight loss ^24^. The differences in the study results might relate to differences in duration and size of the two studies. Leptin and adiponectin are adipokines with known and opposing effects on adaptive and innate immune cells ^27^. Leptin promotes T-cell proliferation and proinflammatory cytokine secretion and inhibits Treg cell induction ^28^ ^29^. Leptin levels are associated with MS risk ^30^, being higher in CSF and serum of pwMS and inversely correlated with circulating Treg numbers ^31^. Conversely, adiponectin promotes Treg induction and function ^32^ and may be reduced in pwMS ^33^. We found serum adiponectin levels to be increased with iCR and this could contribute to its anti-inflammatory effects.

Epidemiological studies have linked a poor diet and obesity with increased disability and disease progression in pwMS ^34^ ^35^, while healthier dietary habits are associated with lower disability ^36^ ^37^. Yet, association studies do not establish causation which could be better evaluated in randomized clinical studies. Preclinical evidence, including from our group, showed that CR ameliorates clinical course and neuropathology of murine experimental autoimmune encephalomyelitis (EAE), through multiple mechanisms including effects on adiposity and the gut microbiome ^6^. This trial in pwMS demonstrates that iCR can modify the immune inflammatory and metabolic profile and antagonize some of the underlying disease pathogenic mechanisms. iCR led to an increase in naïve CD4+ T cells and in Tregs and reduction of Teffs. Notably, no changes in total leukocyte counts were observed with iCR, contrary to what is seen with long-term daily CR ^38^ ^39^, and no significant effect was observed on myeloid cell subsets. These results align with the previous study by Fitzgerald in pwMS ^40^ and with immunomodulatory effects attributed to CR in humans ^38^. Tregs can modulate immune tolerance by detecting nutritional changes and energy intake ^41^. In this study, Tregs from iCR group were more metabolically active than Tregs from the control group while no metabolic changes were observed in the Teff cell compartment. These results are suggesting that iCR can promote tolerogenic Treg cell metabolism.

Beneficial effects of CR include improved vascular and metabolic risk factors which are associated with worse MS outcomes ^42^. Our 12-week study showed no major improvement of the clinical lipid profile or glucose tolerance. However, deeper lipidomic profiling revealed increases in LPC, LPE and PI species after iCR. Lysophospholipids are metabolic intermediates generated via the hydrolysis of membrane or lipoprotein-associated phospholipids by phospholipases. The profile of LPC and LPE species increased in response to iCR reflects the most abundant fatty acids in phospholipids (16:0, 18:0 18:1, 18:2, 20:4) and may result from fasting-dependent lipolysis of adipocyte phospholipids, in line with the reduced fat mass. It is unlikely to result from lipoprotein-associated phospholipase A_2_ activity, since this activity is positively correlated with caloric intake ^43^. Recent work has shown that circulating LPCs are inversely associated with BMI ^44^ ^45^, inflammatory markers including C-reactive protein ^45^ ^46^ and cardiovascular disease risk ^45^. Importantly, transport of polyunsaturated LPCs across the blood brain barrier is essential for brain development and myelin synthesis ^47^ ^48^. iCR decreased levels of HexCer and LacCer in iCR compared to control groups may reflect an overall reduction in sphingolipid synthesis associated with improved metabolic health, since these sphingolipids have been associated with increased cardiovascular risk in some ^49^, but not all studies ^50^. LacCer synthesis in astrocytes may promote neuroinflammation in EAE ^51^. Future studies should be done to determine if iCR affects LacCer synthesis in the CNS.

This study revealed several beneficial effects of iCR in people with RRMS. Limitations of this study include its small patient numbers and that it was only 12 weeks in duration. Even so, we observed significant effects on some objective laboratory variables within 12 weeks. The low subject numbers could limit the power to detect small-size effects. The enrolled subjects were not blinded to the intervention, which could influence some of the secondary outcomes. The intrinsic limitation of non-blinding was somewhat mitigated by blinded assessments of laboratory outcomes, including the primary outcome of leptin levels. Another limitation is that the study enrolled only people with RRMS, and results will need validation in people with progressive disease.

In summary, calorie restriction has the potential to positively impact MS disease outcomes, but the rigorous study of diet interventions is difficult. This randomized study utilized quantitative and objective measures of changes in adipokines and other blood factors, along with subjective clinical measures, to assess effects of iCR in MS patients. The objective beneficial results from the present study along with strong preclinical data supporting CR anti-inflammatory and neuroprotective effects provide strong rationale for continued study of iCR in MS. Larger and longer studies of CR and iCR in pwMS, including blinded imaging outcomes for all subjects, are warranted.

## Data Availability

All data produced in the present study are available upon reasonable request to the authors.

## ACKNOWLEDGMENT

The authors thank Courtney Dula (main study coordinator), and the neurologists, MS fellows, and additional study coordinators at the John L. Trotter MS Center at WUSM for their assistance with patient enrollment, study assessments, and other aspects of this study. Most importantly, the authors thank the people living with MS who participated and made this work possible.

This work is dedicated to the memory of Dr. Jennifer Stark, who made significant contributions to the preclinical studies that were fundamental to the basis of this clinical trial. She was a dear friend and colleague, and she will continue to live in our hearts.

## STUDY FUNDING

This study was funded by the National MS Society (grant # RG-1607-25158 to L.P.). L.G was supported during this study by Fondazione Italiana Sclerosi Multipla (FISM) (2018/B/1) and by the National Multiple Sclerosis (NMSS) Postdoctoral fellowship (FG-1907-34474). CC was supported by the National Multiple Sclerosis Society (TA-1805– 31003) and the Department of Defense (MS200066). L.F. was supported by the Bakewell Foundation during this study. Some of the assessments were supported by EU funding from Fondazione Italiana Sclerosi Multipla (FISM no. 2018/S/5) to G.M.; the MUR PNRR Extended Partnership (INF-ACT no. PE00000007 and MNESYS no. PE00000006) to G.M. and PNRR-Salute (no. PNRR-MAD-2022-12375634) to G.M. A.H.C was supported by the Manny & Rosalyn Rosenthal-Dr. John L. Trotter MS Center Chair in Neuroimmunology of Barnes-Jewish Hospital Foundation.

## COMPETING INTERESTS

L.P. has received research funding from the National MS Society, the NIH, the Department of Defense and Fondazione Italiana Sclerosi Multipla; she has been funded by Alector and Biogen for a project not related to the one included in this manuscript. She is one of the Editor-in Chief of Journal of Neuroimmunology. AHC received compensation for consulting for Biogen, EMD Serono, Bristol Myers Squibb, TG Therapeutics, Octave, Genentech, Roche, Novartis, Horizon and Janssen (J&J). AHC was supported by the Manny & Rosalyn Rosenthal-Dr. John L. Trotter MS Center Chair in Neuroimmunology during this study. GFW received compensation for consulting for EMD Serono, Genzyme, Novartis, Sangamo, Roche, Alumis, and the US Department of Justice. He has received research grant funding from the NIH, National MS Society, Doris Duke Foundation, US Department of Veterans Affairs, Biogen, EMD Serono, and Genentech. He serves on the editorial boards of Neurology: Neuroimmunology & Neuroinflammation and the Journal of Neuroimmunology. He serves on advisory boards for Progentec and Genentech. RTN has consulted for Alexion Pharmaceuticals, Biogen, Bristol Myers Squibb, Celltrion, Genentech, Genzyme, EMD Serono, Horizon Therapeutics, Novartis, TG Therapeutics. Amber Salter receives research funding from Multiple Sclerosis Society of Canada, National Multiple Sclerosis Society, CMSC and the Department of Defense Congressionally Directed Medical Research Program and is a member of editorial board for Neurology. She serves as a consultant for Gryphon Bio, LLC and Abata Therapeutics, LLC. She is a member of the Data and Safety Monitoring Board for Premature Infants Receiving Milking or Delayed Cord Clamping (PREMOD2), Central Vein Sign: A Diagnostic Biomarker in Multiple Sclerosis (CAVS-MS), Ocrelizumab for Preventing Clinical Multiple Sclerosis in Individuals With Radiologically Isolated Disease (CELLO) and Methotrexate treatment of Arthritis caused by Chikungunya virus (MARCH). She holds the Kenney Marie Dixon-Pickens Distinguished Professorship in Multiple Sclerosis Research.

---

**eFIGURE 1.**
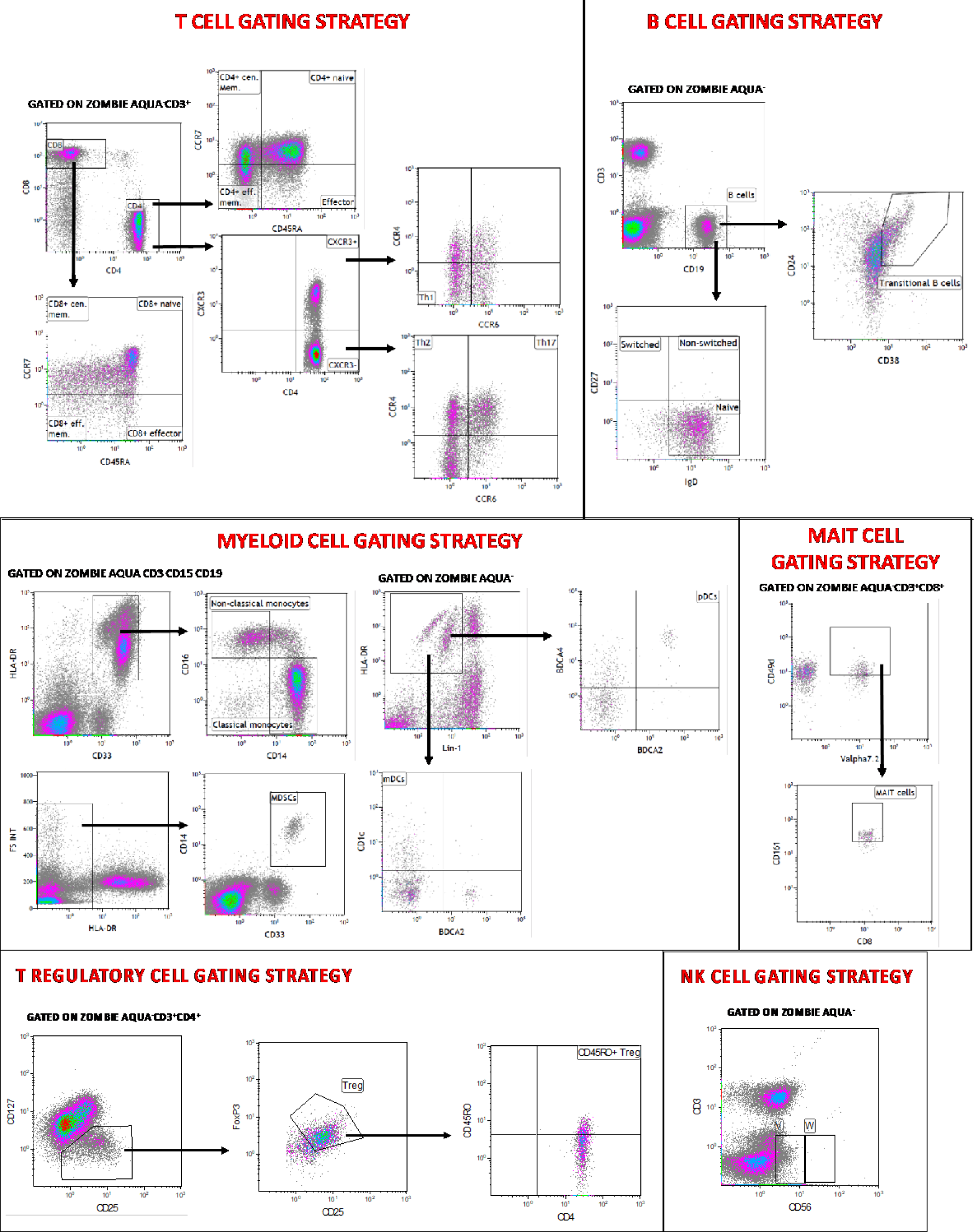
Gating strategy used to analyze T, B, NK and myeloid cells

**eFIGURE 2.**
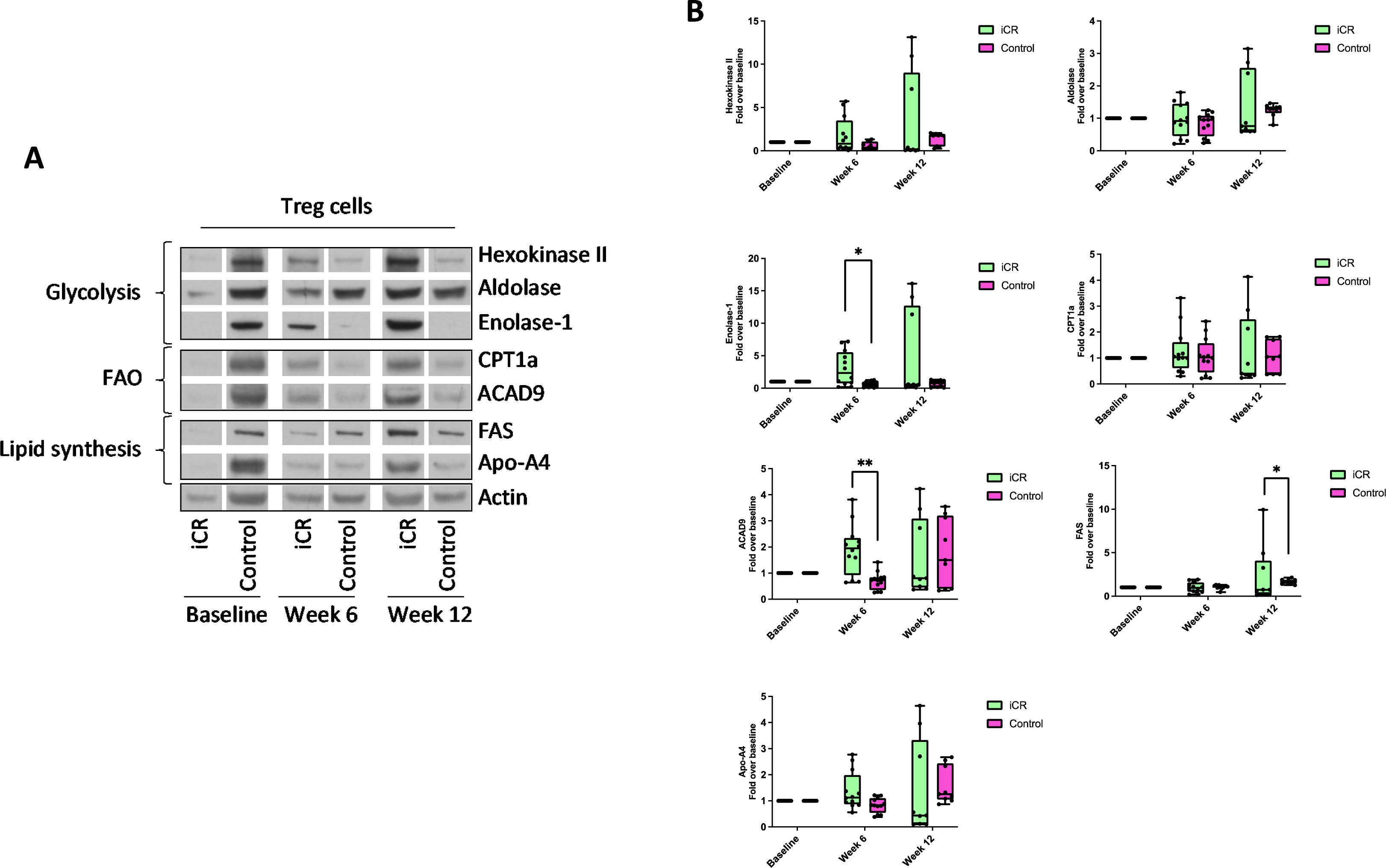
(A) Western blot analysis of Hexokinase II, Aldolase, Enolase-1, CPT1a, ACAD9, FAS and Apo-A4 in Treg cells from iCR and Control groups, at baseline, weeks 6 and weeks 12 of dietary regimen. (B) Densitometric quantification of the specific protein products normalized on actin, and presented as fold over baseline of iCR and Control groups, respectively. Data are from n=2 independent experiments from 4 iCR and Control subjects. Statistical analysis was performed by using Kruskal-Wallis test (mean±sem); *p<0.05; **p<0.01.

**eFIGURE 3.**
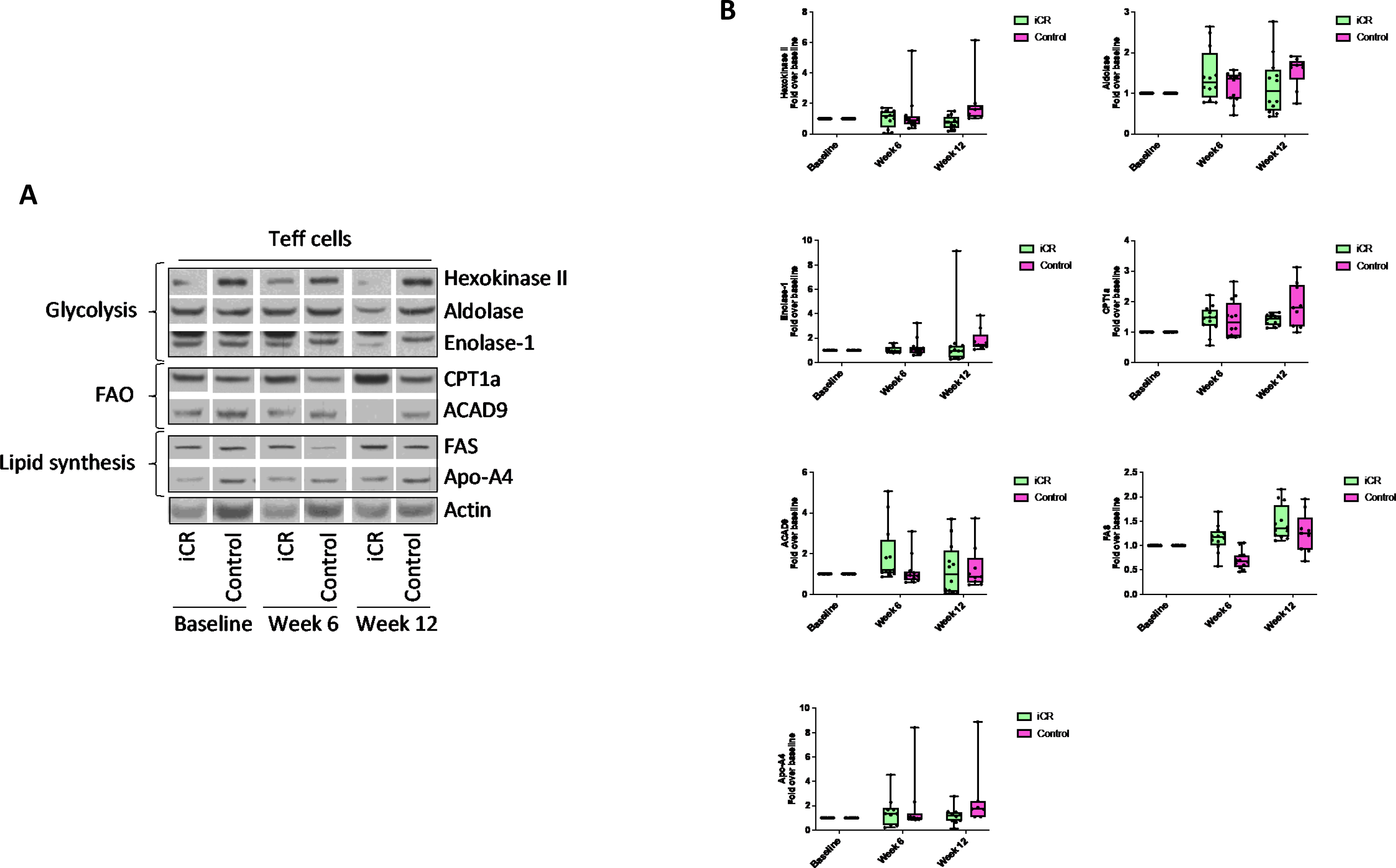
(A) Western blot analysis of Hexokinase II, Aldolase, Enolase-1, CPT1a, ACAD9, FAS and Apo-A4 in Teffs cells from iCR and Control groups, at baseline, weeks 6 and weeks 12 of dietary regimen. (B) Densitometric quantification of the specific protein products normalized on actin, and presented as fold over baseline of iCR and Control groups, respectively. Data are from n=2 independent experiments from 4 iCR and Control subjects. Statistical analysis was performed by using Kruskal-Wallis test (mean±sem).

**eFIGURE 4.**
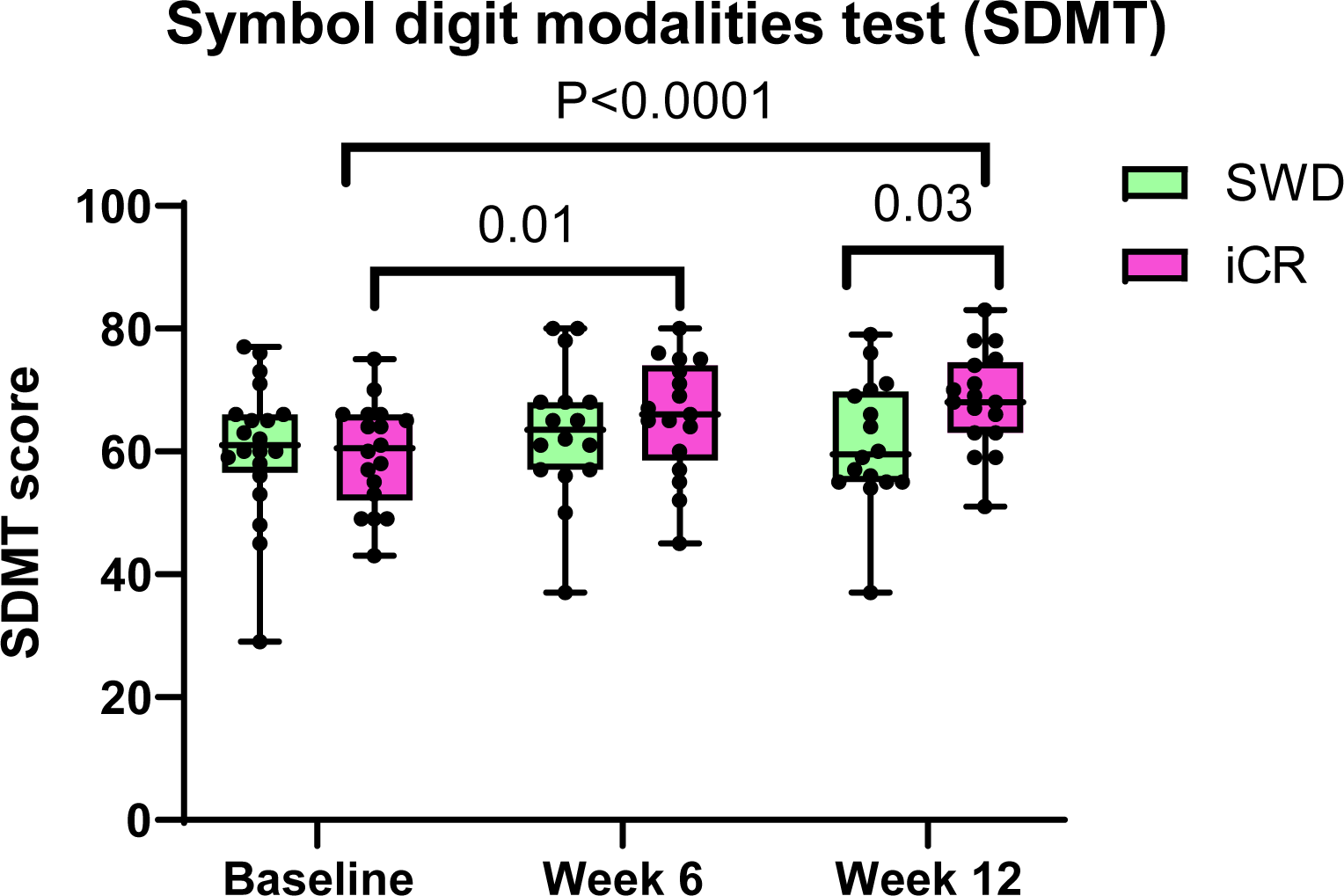
Changes in cognitive outcomes in the iCR and Control groups over the course of the study. Participants randomized to the iCR group showed a significant increase in the SDMT score between baseline, week 6 and week 12. At week 12 the SDMT score was significantly higher in the iCR group compared to the Control group. Individual values for each subject are reported in the graph. The box extends from the 25th to 75th percentiles. Horizontal bars are median and error bars are min and max values. SDMT=Symbol Digit Modality Test.

## Notes

### Clinical Trial

The study was registered at ClinicalTrials.gov (NCT03539094).

### Author Declarations

The study was approved by the Washington University School of Medicine Institutional Review Board (#201707010). Written informed consent was signed by all participants.

