## Supplementary material for "RANDOMIZED CONTROLLED TRIAL OF INTERMITTENT CALORIE RESTRICTION IN PEOPLE WITH MULTIPLE SCLEROSIS": E-methods

**Supplementary Information**

**RESEARCH DESIGN AND METHODS**

***Adipokines and other analytes in blood.*** High sensitivity C-reactive protein (hsCRP) testing was performed using a particle enhanced immunoturbidimetric essay employing Roche reagents analyzed on the Roche cobas c501. Cortisol, insulin and IGF-1 were measured by electrochemiluminescence using Roche Elecsys kits on the Roche cobas e601; β-hydroybutyrate was measured by an enzymatic assay using Stanbio reagents (Sekisui Diagnostics) on the Roche cobas c501. Commercial ELISA Quantikine kits (R&D Systems) were used to measure high molecular weight (HMW) adiponectin and IL-6; leptin was analyzed by radioimmunoassay using EMD Millipore kits.  Plasma neurofilament light chains (pNfL) were measured using the plasma NfL assay by Quanterix. Serum lipids, complete metabolic panel (CMP), cell blood count (CBC), and urinalysis were performed by the Core Laboratory on blood and urine collected at baseline, weeks 6 and 12.

***Oral glucose tolerance test (OGTT)*.** Plasma glucose (glucose oxidase method, Stat Plus, Yellow Springs Instruments Co, Yellow Springs, OH) and insulin (ECLIA electrochemiluminescence, Elecsys Roche Diagnostic, Lewes England, on the Roche cobas e601) were measured in blood samples were drawn at baseline (fasting) and 30, 60, 90 and 120 minutes after the beverage ingestion. Insulin sensitivity and gluco-metabolism were calculated using the homeostatic model assessment method ^21^.
