## Supplementary material for "RANDOMIZED CONTROLLED TRIAL OF INTERMITTENT CALORIE RESTRICTION IN PEOPLE WITH MULTIPLE SCLEROSIS": Supp. Table 1

| **eTable 1. HEI 2015 total and component scores for dietary intake at baseline, week 6 and 12** | | | | | | | | | | |
| --- | --- | --- | --- | --- | --- | --- | --- | --- | --- | --- |
| **Factor** |  | **Baseline** | | | **Week 6 ^1^** | | | **Week 12 ^2^** | | |
|  | **Score Range^** | **Control (N=20)** | **iCR (N=22)** | **p-value** | **Control  (N=18)** | **iCR  (N=19)** | **p-value** | **Control  (N=18)** | **iCR  (N=19)** | **p-value** |
| **HEI 2015 Total Score** | 0-100 | 51.9±7.1 | 51.9±10.0 | 0.99 | 50.6±8.0 | 48.7±8.6 | 0.53 | 50.2±12.5 | 49.1±10.3 | 0.79 |
| HEI 2015 Total Fruits | 0-5 | 1.3±1.01 | 2.1±1.6 | 0.069 | 0.82±0.88 | 1.3±1.08 | 0.13 | 1.2±1.09 | 1.3±1.4 | 0.85 |
| HEI 2015 Whole Fruits | 0-5 | 1.6±1.3 | 2.6±1.8 | ***0.043*** | 1.3±1.2 | 1.5±1.3 | 0.58 | 1.5±1.3 | 1.4±1.4 | 0.79 |
| HEI 2015 Total Vegetables | 0-5 | 3.4±0.86 | 3.3±0.78 | 0.47 | 3.7±0.85 | 3.3±0.87 | 0.14 | 4.2±0.82 | 3.2±1.00 | ***0.007*** |
| HEI 2015 Greens and Beans | 0-5 | 2.8±1.5 | 2.5±1.5 | 0.44 | 2.8±1.5 | 2.1±1.4 | 0.17 | 2.9±1.4 | 2.2±1.4 | 0.13 |
| HEI 2015 Whole Grains | 0-10 | 3.4±2.6 | 2.8±2.5 | 0.42 | 3.0±2.1 | 2.2±1.7 | 0.24 | 2.5±2.3 | 2.2±2.1 | 0.62 |
| HEI 2015 Dairy | 0-10 | 4.2±2.6 | 4.5±2.4 | 0.75 | 4.8±2.6 | 3.6±1.6 | 0.11 | 4.4±2.6 | 4.0±1.5 | 0.60 |
| HEI 2015 Total Protein Foods | 0-5 | 4.5±0.65 | 4.6±0.54 | 0.34 | 4.5±0.65 | 3.3±0.49 | ***<0.001*** | 4.6±0.70 | 3.3±0.56 | ***<0.001*** |
| HEI 2015 Seafood and Plant Proteins | 0-5 | 2.8±1.9 | 3.0±1.4 | 0.64 | 2.7±1.5 | 1.5±1.1 | ***0.010*** | 2.5±1.7 | 2.0±1.4 | 0.30 |
| HEI 2015 Fatty Acids | 0-10 | 5.3±2.3 | 4.3±2.4 | 0.17 | 5.2±2.5 | 5.3±1.7 | 0.86 | 5.8±2.8 | 5.4±1.7 | 0.66 |
| HEI 2015 Refined Grains | 0-10 | 6.3±1.9 | 6.8±2.0 | 0.47 | 7.0±2.1 | 7.2±2.0 | 0.81 | 5.4±2.8 | 6.9±1.9 | 0.075 |
| HEI 2015 Sodium | 0-10 | 3.9±1.9 | 4.1±2.5 | 0.87 | 3.3±2.3 | 3.6±2.5 | 0.69 | 2.6±2.1 | 3.4±2.1 | 0.30 |
| HEI 2015 Added Sugars | 0-10 | 7.8±2.3 | 7.3±2.7 | 0.58 | 7.4±2.6 | 8.3±1.7 | 0.25 | 8.4±2.0 | 8.0±2.2 | 0.56 |
| HEI 2015 Saturated Fats | 0-10 | 4.5±2.4 | 4.1±2.7 | 0.63 | 4.0±2.8 | 5.4±1.6 | 0.084 | 4.1±2.5 | 5.9±1.5 | ***0.023*** |
| HEI: Healthy Eating Index, calculated by NDSR as a measure of diet quality. A higher HEI total score indicates a diet that aligns better with the recommendations. ^Meets Recommendations of the Dietary Guidelines for Americans 2015-2020. Values presented as Mean ± SD. p-values obtained by ANOVA. ^1^Data not available for all subjects. Missing values for all scores: 3. ^2^ Data not available for all subjects. Missing values for all scores: 4 | | | | | | | | | | |
