## Supplementary material for "RANDOMIZED CONTROLLED TRIAL OF INTERMITTENT CALORIE RESTRICTION IN PEOPLE WITH MULTIPLE SCLEROSIS": Supp. Table 2

| **eTable 2: List of reagents for flow cytometry, western blot and lipidomic** | | | | |
| --- | --- | --- | --- | --- |
| **eTable 2 (a).** List of antibodies used in flow cytometry studies | | | | |
| **Reagent or Resources** | | **Supplier/Brand** | **Clone** | |
| Anti-human CD14 | | BD Biosciences | M0P9 | |
| Anti-human CD16 | | BioLegend | 3G8 | |
| Anti-human CD161 | | BioLegend | HP-3G10 | |
| Anti-human CD183(CXCR3) | | BioLegend | G025H7 | |
| Anti-human CD19 | | BioLegend | HIB19 | |
| Anti-human CD194 (CCR4) | | BD Biosciences | 1G1 | |
| Anti-human CD196 (CCR6) | | BD Biosciences | 11A9 | |
| Anti-human CD197 (CCR7) | | BD biosciences | 150503 | |
| Anti-human CD1c | | BioLegend | L161 | |
| Anti-human CD25 | | BioLegend | BC96 | |
| Anti-human CD27 | | BD Biosciences | L128 | |
| Anti-human CD3 | | BioLegend | HIT3a | |
| Anti-human CD303 (BDCA2) | | BioLegend | 201A | |
| Anti-human CD304 (BDCA4) | | BioLegend | 12C2 | |
| Anti-human CD33 | | BioLegend | WM53 | |
| Anti-human CD38 | | BioLegend | HB-7 | |
| Anti-human CD39 | | BioLegend | A1 | |
| Anti-human CD4 | | Beckman Coulter | 13B8.2 | |
| Anti-human CD45RA | | BD Biosciences | HI100 | |
| Anti-human CD45RO | | BD Biosciences | UCHL1 | |
| Anti-human CD49d | | BioLegend | 9F10 | |
| Anti-human CD56 | | BioLegend | 5.1H11 | |
| Anti-human CD62L | | BD Biosciences | DREG-56 | |
| Anti-human CD69 | | BioLegend | FN50 | |
| Anti-human CD8 | | BioLegend | SK1 | |
| Anti-human CD86 | | BioLegend | IT2.2 | |
| Anti-human Foxp3 | | BioLegend | 259D | |
| Anti-human HLA-DR | | Beckman Coulter | B8.12.2 | |
| Anti-human Lineage Cocktail 1 (lin 1) (CD3, CD14, CD16, CD20, CD56) | | BioLegend | UCHT1 | |
| Anti-human Va7.2 | | BioLegend | 3C10 | |
| Zombie Aqua Fixable viability dye | | BioLegend |  | |
| **eTable 2 (b).** List of antibodies used in Western Blot | | | | |
| **Reagent or Resources** | **Supplier/Brand** | | | **Dilutions** |
| ACAD9 | Cell Signaling technology | | | 1:1000 |
| Actin probing antibody | Santa Cruz Biotechnology | | | 1:1000 |
| Aldolase | Cell Signaling technology | | | 1:1000 |
| Apo-A4 | Cell Signaling technology | | | 1:1000 |
| CPT1a | Cell Signaling technology | | | 1:1000 |
| Enolase-1 | Cell Signaling technology | | | 1:1000 |
| FAS | Cell Signaling technology | | | 1:1000 |
| Hexokinase II | Cell Signaling technology | | | 1:1000 |

| **eTable 2 (c).** List of internal standards used for lipidomics | | | |
| --- | --- | --- | --- |
| **Reagent or Resources** | **Supplier/Brand** | **Catalogue number** | **Amount per sample (nmol)** |
| PC(19:0/19:0) | Avanti Polar Lipids (Sigma Aldrich) | 850367P | 5 |
| SM(d18:1/17:0) | Cayman Chemical | 25592 | 2 |
| GluCer(d18:1/17:0) | Avanti Polar Lipids (Sigma Aldrich) | 860569P | 2 |
| PS(17:0/17:0) | Avanti Polar Lipids (Sigma Aldrich) | 840028 | 2 |
| PE(17:0/17:0) | Avanti Polar Lipids (Sigma Aldrich) | 830756 | 2 |
| PG(17:0/17:0) | Avanti Polar Lipids (Sigma Aldrich) | 830456 | 2 |
| PA(17:0/17:0) | Avanti Polar Lipids (Sigma Aldrich) | 830856 | 1 |
| PI(d7-18:1/15:0) | Avanti Polar Lipids (Sigma Aldrich) | 791641C | 1 |
| Cer(d18:1/17:0) | Avanti Polar Lipids (Sigma Aldrich) | 860517P | 0.5 |
| LacCer(d18:1/12:0) | Avanti Polar Lipids (Sigma Aldrich) | 860545P | 0.5 |
| LPC(17:0) | Avanti Polar Lipids (Sigma Aldrich) | 855676 | 0.5 |
| DG(d7-18:1/15:0) | Avanti Polar Lipids (Sigma Aldrich) | 791647C | 0.5 |
| LPE(17:1) | Avanti Polar Lipids (Sigma Aldrich) | 856707 | 0.5 |
| LPS(17:1) | Avanti Polar Lipids (Sigma Aldrich) | 858141 | 0.5 |
| Sph(d17:1) | Avanti Polar Lipids (Sigma Aldrich) | 860640P | 0.2 |
| S1P(d17:1) | Cayman Chemical | 22498 | 0.2 |
| AcCa(d3-16:0) | Avanti Polar Lipids (Sigma Aldrich) | 55107 | 0.2 |
