## Supplementary material for "RANDOMIZED CONTROLLED TRIAL OF INTERMITTENT CALORIE RESTRICTION IN PEOPLE WITH MULTIPLE SCLEROSIS": Supp. Table 3

|  | **Baseline** | | | **Week 6** | | | **Week 12** | | |
| --- | --- | --- | --- | --- | --- | --- | --- | --- | --- |
| **Factor** | **Control (N=20)** | **iCR (N=22)** | **p-value** | **Control (N=17)** | **iCR (N=17)** | **p-value** | **Control* (N=16)** | **iCR (N=17)** | **p-value** |
| **HEI Total Score** | 51.9±7.1 | 51.9±10.0 | 0.99 | 50.6±8.0 | 48.7±8.6 | 0.53 | 50.2±12.5 | 49.1±10.3 | 0.79 |
| **Energy(kcals)** | 2131.9±319.6 | 2034.4±308.1 | 0.32 | 1996.9±387 | 1643.3±390 | ***0.012*** | 1771.1±249 | 1603.2±399 | 0.16 |
| **Calories from Fat** (%) | 39.5±5.2 | 39.3±7.6 | 0.91 | 42.1±7.0 | 39.6±6.2 | 0.27 | 41.7±4.7 | 35.5±7.1 | ***0.006*** |
| **Calories from Carbohydrates** (%) | 41.3±8.5 | 41.8±9.6 | 0.86 | 39.8±7.7 | 43.3±7.5 | 0.19 | 40.9±6.4 | 45.9±7.6 | ***0.047*** |
| **Calories from Protein** (%) | 15.7±3.2 | 16.7±3.0 | 0.31 | 16.2±2.8 | 14.3±2.7 | 0.059 | 16.3±3.3 | 16.2±4.4 | 0.97 |
| **Calories from Saturated Fatty Acids** (%) | 12.8±2.9 | 13.3±3.3 | 0.60 | 14.0±3.8 | 11.8±2.5 | 0.058 | 13.1±2.5 | 10.7±1.9 | ***0.004*** |
| **Total Protein** (gr) | 81.4±16.7 | 84.2±18.6 | 0.62 | 78.0±16.3 | 64.6±15.5 | ***0.019*** | 71.3±11.6 | 65.1±17.2 | 0.24 |
| **Animal Protein** (gr) | 54.2±19.0 | 57.6±17.9 | 0.55 | 54.0±13.5 | 42.3±13.1 | ***0.016*** | 46.9±13.7 | 42.3±15.6 | 0.38 |
| **Plant Protein** (gr) | 27.3±9.0 | 26.6±8.4 | 0.80 | 24.1±8.4 | 22.3±6.5 | 0.50 | 24.4±8.4 | 22.8±6.7 | 0.55 |
| **Total Fruit Servings^Ψ^** | 0.93±0.76 | 1.6±1.4 | 0.071 | 0.50±0.55 | 0.88±0.94 | 0.16 | 0.83±0.86 | 0.99±1.08 | 0.64 |
| **Total Vegetable Servings^Ψ^** | 4.3±1.9 | 3.6±1.7 | 0.24 | 4.2±1.7 | 4.9±2.7 | 0.35 | 4.4±1.8 | 4.2±2.1 | 0.79 |
| **Whole Grain Servings^Ψ^** | 1.07±1.03 | 0.83±0.99 | 0.44 | 0.77±0.85 | 0.55±0.49 | 0.36 | 0.63±0.70 | 0.58±0.67 | 0.84 |
| **Calorie restriction in iCR** (%)**^#^** | NA | NA | NA |  | 21.5±14.4 |  |  | 23.1±14.1 |  |

**eTable 3: Characteristics of dietary intake at baseline, week 6 and 12 in the iCR and Control groups.**

Data for baseline, week 6 and 12 are the average of a 4-day food diary (2 weekdays and weekends). For the iCR group the 4-day food diary contains 1 fast day and 3 non-fast days. For the Control group the 4-day food diary contains 4 days of their usual intake. Values presented as Mean ± SD. p-values calculated by ANOVA. Intention to treat analysis was performed; HEI: Healthy Eating Index, calculated by NDSR as a measure of diet quality; iCR: intermittent calorie restriction; *One subject was missing food diary information at week 12; ^Ψ^Serving sizes have been assigned to each food based on the recommendations made by the 2000 Dietary Guidelines for Americans when available; for foods not included in recommendations (e.g., cookies, fruit drinks), Food and Drug Administration (FDA) serving sizes from 1993 have been used.^#^ Calorie restriction at week 6 (or 12) was calculated as [(Baseline_Energy_kcal -W6_Energy_kcal)/Baseline_Energy_kcal]*100.
