## Supplementary material for "RANDOMIZED CONTROLLED TRIAL OF INTERMITTENT CALORIE RESTRICTION IN PEOPLE WITH MULTIPLE SCLEROSIS": Supp. Table 4

|  |  | **Intermittent Caloric Restriction (iCR)** | | **Control** | | **iCR vs Control** | | |
| --- | --- | --- | --- | --- | --- | --- | --- | --- |
|  |  | **Mean Change**  **(95% CI)** | ***P*-value** | **Mean Change**  **(95% CI)** | ***P*-value** |  | **Mean Difference**  **(95% CI)** | ***P*-value** |
| **LIPID PANEL** | | | | | | | | |
| **Total cholesterol** (mg/dL) | ***0-6 weeks*** | -0.98  (-10.09, 8.13) | 0.96 | 1.76  (-7.4, 10.92) | 0.88 | ***Week 6*** | -0.67  (-20.83, 19.5) | 0.97 |
|  | ***0-12 weeks*** | -1.28  (-10.39, 7.84) | 0.83 | 4.67  (-4.48, 13.83) | 0.25 | ***Week 12*** | 2.54  (-17.62, 22.71) | 0.62 |
| **LDL-cholesterol** (mg/dL) | ***0-6 weeks*** | 0.63  (-7.41, 8.67) | 0.73 | 1.54  (-6.55, 9.62) | 0.98 | ***Week 6*** | -2.76  (-19.27, 13.76) | 0.89 |
|  | ***0-12 weeks*** | 1.16  (-6.88, 9.2) | 0.73 | 2.38  (-5.71, 10.46) | 0.50 | ***Week 12*** | -2.45  (-18.96, 14.06) | 0.84 |
| **HDL-cholesterol** (mg/dL) | ***0-6 weeks*** | -5.91  (-8.52, -3.3) | ***<0.0001*** | -2.24  (-4.86, 0.37) | 0.07 | ***Week 6*** | 2.57  (-5.8, 10.95) | 0.84 |
|  | ***0-12 weeks*** | -2.32  (-4.94, 0.29) | ***0.04*** | -0.4  (-3.02, 2.22) | 0.69 | ***Week 12*** | 0.83  (-7.55, 9.2) | 0.93 |
| **Non-HDL Cholesterol** (mg/dL) | ***0-6 weeks*** | 5.09  (-3.45, 13.63) | 0.27 | 3.637  (-4.94, 12.21) | 0.35 | ***Week 6*** | -3.742  (-24.04, 16.55) | 0.17 |
|  | ***0-12 weeks*** | 1.208  (-7.33, 9.74) | 0.83 | 4.725  (-3.85, 13.3) | 0.23 | ***Week 12*** | 1.229  (-19.07, 21.52) | 0.31 |
| **Triglycerides** (mg/dL) | ***0-6 weeks*** | 23.94  (6.42, 41.47) | ***0.01*** | 11.93  (-5.69, 29.55) | 0.12 | ***Week 6*** | -5.67  (-43.12, 31.77) | 0.47 |
|  | ***0-12 weeks*** | 2.06  (-15.46, 19.58) | 0.98 | 13.8  (-3.82, 31.42) | 0.10 | ***Week 12*** | 18.08  (-19.37, 55.52) | 0.56 |
| **Total cholesterol to HDL ratio** | ***0-6 weeks*** | 0.42  (0.19, 0.65) | ***<0.001*** | 0.19  (-0.04, 0.42) | 0.09 | ***Week 6*** | -0.06  (-0.72, 0.6) | 0.63 |
|  | ***0-12 weeks*** | 0.2  (-0.03, 0.43) | 0.11 | 0.13  (-0.10, 0.36) | 0.24 | ***Week 12*** | 0.092  (-0.57, 0.75) | 0.86 |
| **Triglycerides to HDL ratio** | ***0-6 weeks*** | 0.9  (0.4, 1.41) | ***0.001*** | 0.31  (-0.2, 0.82) | 0.20 | ***Week 6*** | -0.07  (-1.14,1) | 0.79 |
|  | ***0-12 weeks*** | 0.33  (-0.19, 0.84) | 0.25 | 0.31  (-0.21, 0.82) | 0.22 | ***Week 12*** | 0.5  (-0.57, 1.57) | 0.76 |
| **GLUCOSE METABOLISM** | | | | | | | | |
| **Fasting glucose** (mg/dL) | ***0-6 weeks*** | -3.48  (-6.82, -0.14) | ***0.05*** | -0.34  (-3.77, 3.1) | 0.95 | ***Week 6*** | 4.76  (-0.64, 10.16) | 0.17 |
|  | ***0-12 weeks*** | 1.23  (-2.11, 4.57) | 0.45 | -0.95  (-4.38, 2.49) | 0.67 | ***Week 12*** | -0.56  (-5.96, 4.85) | 0.51 |
| **Fasting insulin** (mIU/mL) | ***0-6 weeks*** | 0.59  (-1.58, 2.76) | 0.96 | -2.61  (-4.84, -0.38) | 0.16 | ***Week 6*** | -2.26  (-6.3, 1.77) | 0.18 |
|  | ***0-12 weeks*** | 1.8  (-0.37, 3.97) | 0.49 | -0.68  (-2.91, 1.55) | 0.48 | ***Week 12*** | -1.55  (-5.57, 2.48) | 0.18 |
| **Homoeostasis model assessment-HOMA** | ***0-12 weeks*** | 0.94  (0.02, 1.85) | ***0.05*** | 0.54  (-0.41, 1.49) | 0.26 | ***Week 12*** | -0.09  (-1.08, 0.91) | 0.68 |
| **Glucose AUC**  (mg x h/ml) | ***0-12 weeks*** | 1005.24  (-135.13, 2145.62) | 0.08 | -385.22  (-1633.69, 863.25) | 0.53 | ***Week 12*** | 101.28  (-1866.42, 2068.97) | 0.92 |
| **URINALYSIS** | | | | | | | | |
| **Urine pH** | ***0-6 weeks*** | 0.25  (-0.14, 0.63) | 0.06 | -0.1  (-0.5, 0.29) | 0.32 | ***Week 6*** | 0.02  (-0.52, 0.57) | 0.09 |
|  | ***0-12 weeks*** | 0.34  (-0.05, 0.72) | ***0.02*** | -0.1  (-0.5, 0.29) | 0.36 | ***Week 12*** | -0.07  (-0.61, 0.48) | 0.06 |
| **Urinary specific gravity** | ***0-6 weeks*** | -0.003  (-0.007, 0) | 0.06 | -0.001  (-0.004, 0.003) | 0.79 | ***Week 6*** | 0.001  (-0.004, 0.006) | 0.48 |
|  | ***0-12 weeks*** | -0.002  (-0.006, 0.001) | 0.18 | 0  (-0.004, 0.004) | 0.86 | ***Week 12*** | 0.001  (-0.004, 0.006) | 0.52 |

Mean changes within group and mean difference between groups are calculated based on the raw means. Changes over time within group were calculated subtracting the values at week 6 or 12 to the values at baseline. Therefore, negative values correspond to an increase. Mean differences were calculated as follows: iCR mean at 6 (or 12) weeks minus control mean at 6 (or 12) weeks. P values are adjusted for age, sex and MS disease modifying therapy use. Mean reductions or increases reported in the table are not adjusted. LDL: low-density lipoprotein; HDL: high-density lipoprotein; AUC: area under the curve.
