## Supplementary material for "RANDOMIZED CONTROLLED TRIAL OF INTERMITTENT CALORIE RESTRICTION IN PEOPLE WITH MULTIPLE SCLEROSIS": Supp. Table 5

|  |  |  | **Intermittent Caloric Restriction (iCR)** | | **Control** | |  | **iCR vs Control** | |
| --- | --- | --- | --- | --- | --- | --- | --- | --- | --- |
|  |  |  | **Mean Change**  **(95% CI)** | ***P*-value** | **Mean Change**  **(95% CI)** | ***P*-value** |  | **Mean Difference**  **(95% CI)** | ***P*-value** |
| **IMMUNOPHENOTYPING BY FLOW CYTOMETRY** | | | | | | | | | |
| **Total CD4^+^ T cells** | *cells/μl* | ***0-6 weeks*** | 160.47  (-415.13, 736.08) | 0.15 | -414.56  (-999.21, 170.09) | 0.48 | ***Week***  ***6*** | -506.87  (-1136.64, 122.9) | 0.42 |
|  |  | ***0-12 weeks*** | 165.84  (-409.76, 741.44) | 0.08 | 105.32  (-490.74, 701.39) | 0.82 | ***Week***  ***12*** | 7.65  (-632.73, 648.03) | 0.20 |
|  | *% CD4^+^in the CD3^+^ gate* | ***0-6 weeks*** | 4.32  (-4.48, 13.11) | 0.51 | -2.3  (-11.18, 6.58) | 0.70 | ***Week***  ***6*** | -3.59  (-15.59, 8.42) | 0.22 |
|  |  | ***0-12 weeks*** | 5.57  (-3.22, 14.36) | 0.12 | 2.5  (-6.58, 11.59) | 0.92 | ***Week***  ***12*** | -0.04  (-12.19, 12.12) | 0.10 |
| **Naïve CD4^+^ T cells** | *cells/μl* | ***0-6 weeks*** | -7.7  (-59.97, 44.57) | 0.90 | 44.67  (-6.75, 96.09) | 0.09 | ***Week***  ***6*** | 76.6  (-16.85, 170.05) | 0.17 |
|  |  | ***0-12 weeks*** | -44.5  (-96.78, 7.77) | 0.71 | 61.01  (8.35, 113.68) | ***0.01*** | ***Week***  ***12*** | 76.6  (-16.85, 170.05) | ***0.02*** |
|  | *% CD45RA^+^ CCR7^+^ in the CD4^+^gate* | ***0-6 weeks*** | -2.28  (-7.2, 2.65) | 0.43 | 3.22  (-1.61, 8.06) | 0.44 | ***Week***  ***6*** | 10.93  (0.39, 21.48) | 0.18 |
|  |  | ***0-12 weeks*** | -3.55  (-8.48, 1.38) | 0.40 | 5.43  (0.47, 10.38) | ***0.03*** | ***Week***  ***12*** | 14.41  (3.81, 25.02) | ***0.04*** |
| **Central Memory CD4^+^ T cells** | *cells/μl* | ***0-6 weeks*** | 13.26  (-25.34, 51.86) | 0.42 | 52.5  (14.37, 90.63) | ***0.04*** | ***Week***  ***6*** | 14.23  (-43.31, 71.77) | 0.91 |
|  |  | ***0-12 weeks*** | -8.46  (-47.05, 30.14) | 0.72 | 56.64  (17.61, 95.67) | ***0.01*** | ***Week***  ***12*** | 40.09  (-18.06, 98.23) | 0.27 |
|  | *% CD45RA^-^CCR7^+^ in the CD4^+^ gate* | ***0-6 weeks*** | 1.48  (-3.23, 6.18) | 0.59 | 1.96  (-2.69, 6.6) | 0.66 | ***Week***  ***6*** | 0.91  (-6.49, 8.31) | 0.84 |
|  |  | ***0-12 weeks*** | -1.03  (-5.74, 3.68) | 0.34 | 3.25  (-1.5, 8.01) | 0.36 | ***Week***  ***12*** | 4.71  (-2.76, 12.19) | 0.36 |
| **Effector Memory CD4^+^ T cells** | *cells/μl* | ***0-6 weeks*** | 49.26  (-26.48, 125.01) | 0.12 | 35.12  (-39.65, 109.9) | 0.24 | ***Week***  ***6*** | -47.26  (-162.88, 68.36) | 0.30 |
|  |  | ***0-12 weeks*** | 38.44  (-37.3, 114.19) | 0.16 | 62.56  (-14, 139.12) | 0.36 | ***Week***  ***12*** | -9  (-125.78, 107.78) | 0.29 |
|  | *% CD45RA^-^CCR7^-^ in the CD4^+^ gate* | ***0-6 weeks*** | 2.42  (-2.45, 7.28) | 0.23 | -2.47  (-7.24, 2.31) | 0.10 | ***Week***  ***6*** | -10.56  (-20.76, -0.36) | ***0.04*** |
|  |  | ***0-12 weeks*** | 1.49  (-3.37, 6.36) | 0.19 | -6.22  (-11.11, -1.33) | ***0.02*** | ***Week***  ***12*** | -13.39  (-23.64, -3.13) | ***0.02*** |
| **Effector CD4^+^ T cells** | *cells/μl* | ***0-6 weeks*** | 48.17  (-20.98, 117.32) | 0.83 | 10.72  (-57.93, 79.37) | 0.92 | ***Week***  ***6*** | -24.06  (-113.9, 65.77) | 0.69 |
|  |  | ***0-12 weeks*** | 54.1  (-15.05, 123.25) | 0.37 | 47.63  (-22.58, 117.84) | 0.67 | ***Week***  ***12*** | 6.92  (-84.11, 97.95) | 0.55 |
|  | *% CD45RA^+^ CCR7^-^ in the CD4^+^ gate* | ***0-6 weeks*** | 29  (-7.14, 65.14) | 0.38 | -2.57  (-38.85, 33.72) | 0.25 | ***Week***  ***6*** | -2.06  (-40.17, 36.05) | 0.37 |
|  |  | ***0-12 weeks*** | 33.77  (-2.37, 69.92) | ***0.03*** | -0.65  (-37.62, 36.31) | 0.37 | ***Week***  ***12*** | -4.92  (-43.68, 33.84) | 0.11 |
| **Th1 cells** | *cells/μl* | ***0-6 weeks*** | 66.71  (15.34, 118.07) | ***0.02*** | 24.56  (-27.47, 76.59) | 0.11 | ***Week***  ***6*** | -13.31  (-75.34, 48.73) | ***0.03*** |
|  |  | ***0-12 weeks*** | 53.51  (2.14, 104.88) | 0.07 | 26.22  (-26.93, 79.38) | 0.55 | ***Week***  ***12*** | 1.55  (-61.43, 64.53) | 0.14 |
|  | *% CXCR3^+^CCR4^-^ CCR6^-^ in the CD4^+^ gate* | ***0-6 weeks*** | 6.39  (-1.93, 14.72) | ***0.04*** | 1.91  (-6.49, 10.31) | 0.15 | ***Week***  ***6*** | -2.27  (-14.26, 9.72) | 0.32 |
|  |  | ***0-12 weeks*** | -1.7  (-10.03, 6.62) | 0.53 | 5.29  (-3.31, 13.88) | 0.08 | ***Week***  ***12*** | 9.2  (-2.93, 21.33) | 0.33 |
| **Th2 cells** | *cells/μl* | ***0-6 weeks*** | -0.75  (-23, 21.5) | 0.46 | 0.51  (-21.98, 22.99) | 0.43 | ***Week 6*** | 13.93  (-15.2, 43.06) | 0.63 |
|  |  | ***0-12 weeks*** | 11.74  (-10.51, 33.99) | 0.42 | -2.2  (-25.2, 20.8) | 0.78 | ***Week 12*** | -1.27  (-30.8, 28.26) | 0.82 |
|  | *% CXCR3^-^CCR4^+^ CCR6^-^ in the CD4^+^ gate* | ***0-6 weeks*** | -3.59  (-8.03, 0.85) | 0.08 | -1.26  (-5.74, 3.21) | 0.78 | ***Week 6*** | 4.96  (-1.76, 11.68) | 0.26 |
|  |  | ***0-12 weeks*** | -0.35  (-4.79, 4.09) | 0.20 | -0.16  (-4.74, 4.42) | 0.37 | ***Week 12*** | 2.82  (-3.97, 9.61) | 0.56 |
| **Th17 cells** | *cells/μl* | ***0-6 weeks*** | -0.98  (-19.41, 17.45) | 0.89 | 11.3  (-7.35, 29.95) | 0.19 | ***Week 6*** | 7.75  (-15.25, 30.75) | 0.92 |
|  |  | ***0-12 weeks*** | -0.98  (-19.41, 17.45) | 0.45 | -0.86  (-19.92, 18.2) | 0.70 | ***Week 12*** | -2.84  (-26.18, 20.5) | 0.09 |
|  | *% CXCR3^-^CCR4^+^ CCR6^+^ in the CD4^+^ gate* | ***0-6 weeks*** | -2.02  (-5.15, 1.11) | 0.15 | -1.44  (-4.58, 1.71) | 0.98 | ***Week 6*** | -1.27  (-6.43, 3.9) | 0.91 |
|  |  | ***0-12 weeks*** | -0.84  (-3.96, 2.29) | 0.71 | -2.7  (-5.92, 0.53) | 0.11 | ***Week 12*** | -3.71  (-8.92, 1.5) | 0.09 |
| **Regulatory T cells** | *cells/μl* | ***0-6 weeks*** | -2.06  (-11.3, 7.19) | 0.23 | 4.97  (-4.33, 14.27) | ***0.05*** | ***Week 6*** | 7.94  (-7.13, 23.01) | 0.18 |
|  |  | ***0-12 weeks*** | -2.9  (-12.14, 6.34) | 0.45 | 3.64  (-5.89, 13.17) | 0.74 | ***Week 12*** | 7.46  (-7.75, 22.66) | 1.00 |
|  | *% CD25^hi^ CD127^lo^ FoxP3^+^ in the CD4^+^ gate* | ***0-6 weeks*** | 0.84  (-11.33, 13) | 1.00 | -3.15  (-15.2, 8.91) | 0.87 | ***Week 6*** | -3.38  (-18.69, 11.92) | 0.99 |
|  |  | ***0-12 weeks*** | 8.31  (-3.61, 20.23) | 0.38 | -6  (-18.33, 6.32) | 0.47 | ***Week 12*** | -13.71  (-29.04, 1.62) | 0.42 |
| **CD45RO^+^ Treg** | *cells/μl* | ***0-6 weeks*** | -4.93  (-9.66, -0.21) | ***0.04*** | 1.07  (-3.72, 5.85) | 0.80 | ***Week 6*** | 4.71  (-0.89, 10.31) | 0.27 |
|  |  | ***0-12 weeks*** | -3.92  (-8.65, 0.8) | 0.17 | 1.96  (-2.93, 6.85) | 0.87 | ***Week 12*** | 4.59  (-1.09, 10.28) | 0.71 |
|  | *% CD25^hi^ CD127^lo^ FoxP3^+^* *CD45RO^+^ in the CD4^+^ gate* | ***0-6 weeks*** | -14.1  (-25.74, -2.46) | ***0.03*** | -4.11  (-15.96, 7.74) | 0.82 | ***Week 6*** | 3.07  (-9.6, 15.74) | 0.46 |
|  |  | ***0-12 weeks*** | -7.4  (-18.84, 4.04) | 0.22 | 3.29  (-8.56, 15.13) | 0.66 | ***Week 12*** | 3.77  (-8.72, 16.26) | 0.64 |
| **CD8^+^ T cells** | *cells/μl* | ***0-6 weeks*** | 47.04  (-1.42, 95.5) | 0.13 | 18.7  (-30.09, 67.49) | ***0.04*** | ***Week 6*** | -62  (-139.07, 15.07) | 0.55 |
|  |  | ***0-12 weeks*** | 25.72  (-22.74, 74.18) | 0.46 | 24.69  (-25.27, 74.65) | 0.25 | ***Week 12*** | -34.69  (-112.51, 43.12) | 0.49 |
|  | *% CD8^+^ in the CD3^+^ gate* | ***0-6 weeks*** | 0.86  (-2.12, 3.84) | 0.58 | 1.02  (-1.97, 4.01) | 0.21 | ***Week 6*** | -5.66  (-11.24, -0.08) | 0.33 |
|  |  | ***0-12 weeks*** | -0.73  (-3.7, 2.25) | 0.82 | 0.47  (-2.59, 3.54) | 0.43 | ***Week 12*** | -4.62  (-10.24, 1) | 0.48 |
| **Naïve CD8^+^ T cells** | *cells/μl* | ***0-6 weeks*** | 8.88  (-12.53, 30.3) | 0.12 | 8.63  (-12.51, 29.76) | 0.66 | ***Week 6*** | -28.68  (-61.63, 4.26) | 0.06 |
|  |  | ***0-12 weeks*** | 2.53  (-18.89, 23.94) | 0.47 | 19.33  (-2.3, 40.97) | 0.22 | ***Week 12*** | -11.62  (-44.89, 21.65) | 0.60 |
|  | *% CD45RA^+^ CCR7^+^ in the CD8^+^ gate* | ***0-6 weeks*** | 3.9  (-1.01, 8.82) | 0.31 | 0.29  (-4.53, 5.1) | 0.99 | ***Week 6*** | -5.21  (-16.84, 6.42) | 0.38 |
|  |  | ***0-12 weeks*** | 3.76  (-1.15, 8.67) | 0.33 | 2.36  (-2.58, 7.29) | 0.65 | ***Week 12*** | -3  (-14.68, 8.68) | 0.58 |
| **Central Memory CD8^+^ T cells** | *cells/μl* | ***0-6 weeks*** | 9.61  (-9.05, 28.26) | 0.42 | -2.92  (-21.42, 15.59) | 0.63 | ***Week 6*** | -13.97  (-38.73, 10.79) | 0.14 |
|  |  | ***0-12 weeks*** | -2.98  (-21.63, 15.68) | 0.53 | -6.12  (-25.05, 12.8) | 0.57 | ***Week 12*** | -4.6  (-29.68, 20.48) | 0.87 |
|  | *% CD45RA^-^ CCR7^+^ in the CD8^+^ gate* | ***0-6 weeks*** | 4.79  (-2.57, 12.16) | 0.31 | -2.87  (-10.14, 4.4) | 0.21 | ***Week 6*** | -2.96  (-14, 8.07) | 0.28 |
|  |  | ***0-12 weeks*** | -1.8  (-9.16, 5.56) | 0.84 | -2.22  (-9.66, 5.23) | 0.31 | ***Week 12*** | 4.28  (-6.87, 15.43) | 0.98 |
| **Effector Memory CD8^+^ T cells** | *cells/μl* | ***0-6 weeks*** | -0.84  (-26.49, 24.81) | 0.24 | 5.77  (-19.42, 30.96) | 0.15 | ***Week 6*** | -17.87  (-68.46, 32.73) | 0.44 |
|  |  | ***0-12 weeks*** | -10.05  (-35.7, 15.6) | 0.90 | 22  (-3.81, 47.81) | 0.08 | ***Week 12*** | 7.57  (-43.33, 58.48) | 0.70 |
|  | *% CD45RA^-^ CCR7^-^ in the CD8^+^ gate* | ***0-6 weeks*** | -3.22  (-11.26, 4.82) | 0.22 | 5.22  (-2.71, 13.16) | 0.21 | ***Week 6*** | 5.49  (-6.78, 17.77) | 0.50 |
|  |  | ***0-12 weeks*** | -5.83  (-13.87, 2.21) | 0.50 | 3.75  (-4.38, 11.87) | 0.13 | ***Week 12*** | 6.63  (-5.77, 19.03) | 0.60 |
| **Effector CD8^+^ T cells** | *cells/μl* | ***0-6 weeks*** | -1.17  (-33.5, 31.16) | 0.27 | -18.09  (-49.94, 13.76) | 0.71 | ***Week 6*** | -31.11  (-84.75, 22.53) | 0.73 |
|  |  | ***0-12 weeks*** | 11.11  (-21.22, 43.44) | 0.29 | 4.69  (-27.93, 37.31) | 0.68 | ***Week 12*** | -20.6  (-74.7, 33.5) | 0.38 |
|  | *% CD45RA^+^ CCR7^-^ in the CD8^+^ gate* | ***0-6 weeks*** | -5.05  (-11.61, 1.51) | 0.57 | -2.84  (-9.28, 3.6) | 0.98 | ***Week 6*** | 2.15  (-11.84, 16.14) | 0.63 |
|  |  | ***0-12 weeks*** | 4.3  (-2.26, 10.87) | 0.25 | -4.31  (-10.91, 2.28) | 0.50 | ***Week 12*** | -8.68  (-22.74, 5.38) | 0.42 |
| **Mucosal Associated Invariant T cells (MAIT)** | *cells/μl* | ***0-6 weeks*** | 0.6  (-4.97, 6.17) | 0.53 | 3.24  (-2.35, 8.83) | ***0.02*** | ***Week 6*** | -3.28  (-14.13, 7.58) | 0.23 |
|  |  | ***0-12 weeks*** | -1.75  (-7.32, 3.82) | 0.13 | 4.22  (-1.51, 9.95) | 0.15 | ***Week 12*** | 0.05  (-10.87, 10.98) | 0.11 |
|  | *% CD8^+^ Vα7.2^+^CD161^hi^ in the CD3^+^ gate* | ***0-6 weeks*** | -2.3  (-10.61, 6.01) | 0.10 | 6.18  (-2.15, 14.51) | 0.66 | ***Week 6*** | 11.64  (-9, 32.28) | ***0.05*** |
|  |  | ***0-12 weeks*** | -10.63  (-18.94, -2.32) | ***0.03*** | 4.38  (-4.16, 12.92) | 0.57 | ***Week 12*** | 18.17  (-2.55, 38.89) | ***0.02*** |
| **B cells** | *cells/μl* | ***0-6 weeks*** | 36.01  (-177.47, 249.5) | 0.47 | 39.14  (-177.82, 256.09) | 0.42 | ***Week 6*** | -13.67  (-243, 215.65) | 0.72 |
|  |  | ***0-12 weeks*** | 20.42  (-193.06, 233.91) | 0.53 | -144.1  (-365.17, 76.96) | 0.18 | ***Week 12*** | -181.32  (-414.54, 51.89) | 0.82 |
|  | *% CD19^+^ in the Live cell gate* | ***0-6 weeks*** | 1.17  (-4.38, 6.72) | 0.30 | 0.92  (-4.72, 6.55) | 0.23 | ***Week 6*** | -0.89  (-7.11, 5.32) | 0.73 |
|  |  | ***0-12 weeks*** | 0.38  (-5.17, 5.92) | 0.56 | -4.02  (-9.77, 1.72) | 0.26 | ***Week 12*** | -5.04  (-11.36, 1.28) | 0.99 |
| **Naïve B cells** | *cells/μl* | ***0-6 weeks*** | 18.88  (-18.37, 56.14) | 0.59 | 34.3  (-3.51, 72.1) | 0.37 | ***Week 6*** | -0.37  (-42.53, 41.79) | 0.67 |
|  |  | ***0-12 weeks*** | 8.06  (-29.2, 45.32) | 0.64 | 54.75  (16.17, 93.32) | ***0.02*** | ***Week 12*** | 30.9  (-11.95, 73.76) | 0.06 |
|  | *% IgD^+^CD27^-^ in the CD19^+^ gate* | ***0-6 weeks*** | 3.56  (-7.5, 14.62) | 0.88 | 4.37  (-6.83, 15.57) | 0.26 | ***Week 6*** | 5.97  (-7.41, 19.36) | 0.10 |
|  |  | ***0-12 weeks*** | -0.41  (-11.47, 10.65) | 0.90 | 13.69  (2.25, 25.13) | 0.13 | ***Week 12*** | 19.26  (5.67, 32.85) | 0.06 |
| **Non-switched memory B cells** | *cells/μl* | ***0-6 weeks*** | 0.55  (-1.1, 2.21) | ***0.04*** | -0.31  (-1.98, 1.36) | 0.45 | ***Week 6*** | -2.29  (-4.62, 0.04) | 0.08 |
|  |  | ***0-12 weeks*** | 0.88  (-0.77, 2.53) | 0.15 | 1.72  (0.02, 3.43) | 0.22 | ***Week 12*** | -0.59  (-2.95, 1.77) | 0.33 |
|  | *% IgD^+^CD27^+^ in the CD19^+^ gate* | ***0-6 weeks*** | -0.29  (-1.44, 0.86) | 0.62 | 0.08  (-1.08, 1.25) | 0.29 | ***Week 6*** | -0.34  (-1.69, 1.02) | 0.82 |
|  |  | ***0-12 weeks*** | 0.2  (-0.95, 1.34) | 0.22 | 0.9  (-0.29, 2.09) | 0.12 | ***Week 12*** | 0  (-1.38, 1.37) | 0.75 |
| **Switched memory B cells** | *cells/μl* | ***0-6 weeks*** | -0.07  (-10.3, 10.16) | 0.88 | 7.85  (-2.54, 18.23) | 0.09 | ***Week 6*** | -1.79  (-13.3, 9.72) | 0.85 |
|  |  | ***0-12 weeks*** | -0.01  (-10.24, 10.22) | 0.56 | 9.54  (-1.05, 20.13) | ***0.02*** | ***Week 12*** | -0.16  (-11.86, 11.54) | 0.63 |
|  | *% IgD^-^CD27^+^ in the CD19^+^ gate* | ***0-6 weeks*** | -0.05  (-0.62, 0.53) | 0.88 | -0.42  (-1, 0.16) | 0.88 | ***Week 6*** | -1.09  (-2.45, 0.28) | 0.72 |
|  |  | ***0-12 weeks*** | -0.07  (-0.65, 0.51) | 0.53 | 0.43  (-0.17, 1.02) | 0.15 | ***Week 12*** | -0.22  (-1.59, 1.16) | 0.91 |
| **Transitional B cells** | *cells/μl* | ***0-6 weeks*** | 8.7  (0.16, 17.24) | 0.41 | 4.7  (-3.92, 13.32) | 0.30 | ***Week 6*** | -3.96  (-15.85, 7.93) | 0.60 |
|  |  | ***0-12 weeks*** | 5.36  (-3.17, 13.9) | 0.83 | 8.98  (0.16, 17.8) | ***0.04*** | ***Week 12*** | 3.66  (-8.37, 15.7) | 0.26 |
|  | *% CD27^hi^ CD38 ^hi^ in the CD19^+^ gate* | ***0-6 weeks*** | 2.09  (-0.66, 4.83) | 0.99 | 0.22  (-2.54, 2.98) | 0.71 | ***Week 6*** | -2.36  (-7.43, 2.71) | 0.75 |
|  |  | ***0-12 weeks*** | 2.74  (0, 5.49) | 0.82 | -0.23  (-3.06, 2.6) | 0.35 | ***Week 12*** | -3.47  (-8.58, 1.64) | 0.76 |
| **Classical monocytes** | *cells/μl* | ***0-6 weeks*** | -25.04  (-64.82, 14.74) | 0.77 | 67.94  (27.82, 108.05) | ***0.05*** | ***Week 6*** | 61.47  (3.41, 119.54) | 0.65 |
|  |  | ***0-12 weeks*** | 25.97  (-13.82, 65.75) | 0.50 | 65.19  (24.13, 106.25) | ***0.03*** | ***Week 12*** | 7.72  (-51, 66.44) | 0.83 |
|  | *% CD33^+^ HLA-DR^+^CD14^+^CD16^-^ in the CD3^-^CD15^-^CD19^-^ gate* | ***0-6 weeks*** | -8.51  (-21.32, 4.29) | 0.26 | 18.91  (5.96, 31.86) | 0.05 | ***Week 6*** | 16.06  (-0.57, 32.7) | 0.52 |
|  |  | ***0-12 weeks*** | -0.46  (-13.27, 12.35) | 0.40 | 13.87  (0.63, 27.11) | ***0.03*** | ***Week 12*** | 2.97  (-13.89, 19.84) | 0.54 |
| **Non-classical monocytes** | *cells/μl* | ***0-6 weeks*** | -5.96  (-12.28, 0.37) | 0.75 | -1.66  (-8.05, 4.73) | 0.47 | ***Week 6*** | 7.61  (-0.82, 16.03) | 0.19 |
|  |  | ***0-12 weeks*** | 4.86  (-1.47, 11.18) | 0.38 | 1.57  (-4.96, 8.11) | 0.56 | ***Week 12*** | 0.02  (-8.51, 8.56) | 0.64 |
|  | *% CD33^+^ HLA-DR^+^CD14^-^CD16^+^ in the CD3^-^CD15^-^CD19^-^ gate* | ***0-6 weeks*** | -1.73  (-4.03, 0.58) | 0.57 | -1.56  (-3.89, 0.77) | 0.91 | ***Week 6*** | 1.8  (-1.13, 4.74) | 0.32 |
|  |  | ***0-12 weeks*** | 0.53  (-1.78, 2.83) | 0.81 | -0.86  (-3.24, 1.52) | 0.73 | ***Week 12*** | 0.25  (-2.72, 3.23) | 0.51 |
| **Myeloid-derived suppressor cells** | *cells/μl* | ***0-6 weeks*** | -0.25  (-2.92, 2.42) | 0.41 | 1.49  (-1.2, 4.19) | 0.25 | ***Week 6*** | -0.62  (-4.55, 3.3) | 0.26 |
|  |  | ***0-12 weeks*** | 0.37  (-2.31, 3.04) | ***0.02*** | 0.63  (-2.12, 3.39) | 0.19 | ***Week 12*** | -2.11  (-6.07, 1.86) | ***0.05*** |
|  | *% HLA-DR^-^CD33^+^ CD14^+^ in the CD3^-^CD15^-^CD19^-^ gate* | ***0-6 weeks*** | 0.11  (-0.74, 0.97) | 0.09 | 0.36  (-0.5, 1.23) | 0.15 | ***Week 6*** | 0.07  (-1.21, 1.36) | 0.33 |
|  |  | ***0-12 weeks*** | -0.03  (-0.88, 0.83) | ***0.02*** | -0.2  (-1.09, 0.68) | 0.17 | ***Week 12*** | -0.35  (-1.65, 0.95) | 0.14 |
| **Plasmacytoid dendritic cells** - pDCs | *cells/μl* | ***0-6 weeks*** | -0.42  (-1.23, 0.4) | 0.51 | 0.13  (-0.69, 0.95) | 0.93 | ***Week 6*** | -0.8  (-2.97, 1.37) | 0.68 |
|  |  | ***0-12 weeks*** | 0.14  (-0.68, 0.95) | 0.72 | 0.23  (-0.61, 1.06) | 0.61 | ***Week 12*** | -1.26  (-3.43, 0.92) | 0.42 |
|  | *% HLA-DR^+^ Lin-1^-^BDCA2^+^ BDCA4^+^ in the live cell gate* | ***0-6 weeks*** | -0.15  (-0.56, 0.26) | 0.55 | -0.12  (-0.53, 0.29) | 0.43 | ***Week 6*** | -0.52  (-1.44, 0.4) | 0.86 |
|  |  | ***0-12 weeks*** | -0.28  (-0.69, 0.12) | 0.33 | -0.13  (-0.55, 0.29) | 0.34 | ***Week 12*** | -0.4  (-1.32, 0.53) | 0.98 |
| **Myeloid dendritic cells** - mDCs | *cells/μl* | ***0-6 weeks*** | -2.25  (-11.21, 6.71) | 0.60 | 3.48  (-5.59, 12.56) | 0.14 | ***Week 6*** | 0.77  (-10.13, 11.67) | 0.53 |
|  |  | ***0-12 weeks*** | -9.04  (-18, -0.08) | 0.50 | 3.26  (-6.01, 12.53) | 0.46 | ***Week 12*** | 7.34  (-3.73, 18.4) | 0.92 |
|  | *% HLA-DR^+^ Lin-1^-^ CD1c^+^ in the live cell gate* | ***0-6 weeks*** | -0.51  (-3, 1.97) | 0.63 | 0.09  (-2.42, 2.6) | 0.69 | ***Week 6*** | -0.81  (-4.3, 2.67) | 0.99 |
|  |  | ***0-12 weeks*** | -3.86  (-6.35, -1.38) | ***0.03*** | 0.05  (-2.52, 2.61) | 0.95 | ***Week 12*** | 2.49  (-1.03, 6.02) | 0.31 |
| **CD56^dim^ NK cells** | *cells/μl* | ***0-6 weeks*** | -9.72  (-27.67, 8.23) | 0.50 | -3.81  (-21.96, 14.35) | 0.28 | ***Week 6*** | 7.64  (-15.1, 30.38) | 0.91 |
|  |  | ***0-12 weeks*** | 5.16  (-12.79, 23.11) | 0.91 | -2.01  (-20.57, 16.56) | 0.72 | ***Week 12*** | -5.44  (-28.5, 17.63) | 0.88 |
|  | *% CD56^low^in the CD3^-^ gate* | ***0-6 weeks*** | -0.6  (-1.24, 0.03) | 0.13 | -0.16  (-0.8, 0.48) | 0.28 | ***Week 6*** | 0.34  (-0.65, 1.34) | 0.68 |
|  |  | ***0-12 weeks*** | -0.06  (-0.7, 0.57) | 0.24 | -0.04  (-0.69, 0.62) | 0.69 | ***Week 12*** | -0.08  (-1.08, 0.93) | 0.51 |
| **CD56^bright^ NK cells** | *cells/μl* | ***0-6 weeks*** | -0.3  (-1.95, 1.35) | 0.74 | 1.1  (-0.57, 2.76) | 0.10 | ***Week 6*** | -0.05  (-2.66, 2.56) | 0.76 |
|  |  | ***0-12 weeks*** | 0.59  (-1.07, 2.24) | 0.62 | 0.64  (-1.06, 2.34) | 0.75 | ***Week 12*** | -0.05  (-2.66, 2.56) | 0.49 |
|  | *% CD56^hi^ in the CD3^-^ gate* | ***0-6 weeks*** | -0.02  (-0.1, 0.05) | 0.74 | 0.03  (-0.04, 0.1) | 0.16 | ***Week 6*** | -0.01  (-0.12, 0.1) | 0.74 |
|  |  | ***0-12 weeks*** | 0.01  (-0.07, 0.08) | 0.91 | 0.05  (-0.02, 0.12) | 0.93 | ***Week 12*** | -0.02  (-0.14, 0.09) | 0.86 |

Mean changes within group and mean difference between groups are calculated based on the raw means. Changes over time within group were calculated subtracting the values at week 6 or 12 to the values at baseline. Therefore, negative values correspond to an increase. Mean differences were calculated as follows: iCR mean at 6 (or 12) weeks minus control mean at 6 (or 12) weeks. P values are adjusted for age, sex and MS disease modifying therapy use. Mean reductions or increases reported in the table are not adjusted. NK: natural killer.
