## Supplementary material for "RANDOMIZED CONTROLLED TRIAL OF INTERMITTENT CALORIE RESTRICTION IN PEOPLE WITH MULTIPLE SCLEROSIS": Supp. Table 7

**e Table 7**. Significant lipids adjusted for FDR in the iCR group (between baseline and 12 months) and between control and iCR groups at 12 weeks.

| ***iCR 0***  ***vs***  ***iCR 12*** | **Lipids** | **Effect** | **Treat** | **W** | **treat** | **w** | **Estimate** | **Lower** | **Upper** | **p-value** | **FDR (q<0.05)** |
| --- | --- | --- | --- | --- | --- | --- | --- | --- | --- | --- | --- |
| 1 | LPC(16:0) | treat*w | IF | 0 | IF | 12 | -14574.1 | -21676.8 | -7471.5 | 0.0002 | 0.0202 |
| 2 | LPE(16:0) | treat*w | IF | 0 | IF | 12 | -258.0 | -384.3 | -131.8 | 0.0003 | 0.0202 |
| 3 | PI(18:1/18:2) | treat*w | IF | 0 | IF | 12 | -641.1 | -967.6 | -314.6 | 0.0004 | 0.0210 |
| 4 | LPC(18:1) | treat*w | IF | 0 | IF | 12 | -3808.6 | -5838.1 | -1779.1 | 0.0006 | 0.0253 |
| 5 | PI(18:1/20:4) | treat*w | IF | 0 | IF | 12 | -210.8 | -331.4 | -90.1 | 0.0013 | 0.0363 |
| 6 | LPC(18:2) | treat*w | IF | 0 | IF | 12 | -8590.6 | -13542.5 | -3638.6 | 0.0014 | 0.0363 |
| 7 | LPC(22:4) | treat*w | IF | 0 | IF | 12 | -93.2 | -148.9 | -37.5 | 0.0019 | 0.0435 |
| 8 | LPE(18:1) | treat*w | IF | 0 | IF | 12 | -311.1 | -503.1 | -119.2 | 0.0025 | 0.0498 |
| 9 | LPC(20:4) | treat*w | IF | 0 | IF | 12 | -1643.1 | -2670.9 | -615.3 | 0.0028 | 0.0498 |
| ***iCR 12***  ***vs***  ***Control 12*** | **Lipids** | **Effect** | **Treat** | **W** | **treat** | **w** | **Estimate** | **Lower** | **Upper** | **p-value** | **FDR (q<0.05)** |
| 1 | PI(16:1/18:1) | treat*w | IF | 12 | SW | 12 | 261.3 | 144.0 | 378.7 | 0.0001 | 0.0146 |
| 2 | HexCer(d18:1/22:1) | treat*w | IF | 12 | SW | 12 | -13.8 | -21.0 | -6.6 | 0.0005 | 0.0292 |
| 3 | PI(18:1/20:4) | treat*w | IF | 12 | SW | 12 | 456.6 | 210.6 | 702.6 | 0.0007 | 0.0292 |
| 4 | LacCer(d18:1/22:1) | treat*w | IF | 12 | SW | 12 | -6.4 | -9.9 | -2.9 | 0.0008 | 0.0292 |
| 5 | LPC(22:4) | treat*w | IF | 12 | SW | 12 | 172.4 | 77.1 | 267.6 | 0.0009 | 0.0292 |
| 6 | LPE(16:0) | treat*w | IF | 12 | SW | 12 | 379.8 | 165.5 | 594.1 | 0.0011 | 0.0297 |
| 7 | PI(16:0/20:4) | treat*w | IF | 12 | SW | 12 | 1464.4 | 618.8 | 2310.0 | 0.0014 | 0.0317 |
| 8 | DG(18:0/18:1) | treat*w | IF | 12 | SW | 12 | 524.7 | 208.9 | 840.5 | 0.0020 | 0.0403 |
| 9 | PI(16:0/18:1) | treat*w | IF | 12 | SW | 12 | 1138.0 | 431.6 | 1844.3 | 0.0026 | 0.0466 |
| 10 | LPE(18:0) | treat*w | IF | 12 | SW | 12 | 594.8 | 218.8 | 970.8 | 0.0031 | 0.0489 |
