## Supplementary material for "RANDOMIZED CONTROLLED TRIAL OF INTERMITTENT CALORIE RESTRICTION IN PEOPLE WITH MULTIPLE SCLEROSIS": Supp. Table 8

**eTable 8.** Correlation analyses conducted between the 16 lipids that were significantly changed after iCR and clinical variables that were significantly affected by iCR.

|  | **T-statistic** | **Degrees of freedom** | **P-value** | **Pearson's correlation coefficient  (95% CI)** |
| --- | --- | --- | --- | --- |
| **BMI** |  |  |  |  |
| LPC(20:4) | 2.65 | 28 | 0.013 | 0.45 (0.10 to 0.70) |
| LPC(22:4) | 2.53 | 28 | 0.017 | 0.43 (0.08 to 0.69) |
| **EDSS** |  |  |  |  |
| LPC(20:4) | 2.41 | 66 | 0.019 | 0.28 (0.05 to 0.49) |
| **Adiponectin** |  |  |  |  |
| HexCer(d18:1/22:1) | 2.65 | 28 | 0.013 | 0.45 (0.10 to 0.70) |
| PI(18:1/18:2) | 2.32 | 28 | 0.028 | 0.40 (0.05 to 0.67) |
| PI(16:1/18:1) | 2.25 | 28 | 0.032 | 0.39 (0.04 to 0.70) |
| **CHDLr** |  |  |  |  |
| DGI(18:0/18:1) | 2.42 | 28 | 0.023 | 0.42 (0.06 to 0.67) |
| HexCer(d18:1/22:1) | -2.33 | 28 | 0.027 | -0.40 (-0.05 to -0.67) |
| **HDL** |  |  |  |  |
| PI(18:1/18:2) | 2.11 | 28 | 0.044 | 0.37 (0.01 to 0.65) |
| **Monocyte %** |  |  |  |  |
| HexCer(d18:1/22:1) | 2.83 | 28 | 0.009 | 0.47 (0.13 to 0.71) |
| LacCer(d18:1/22:1) | 2.23 | 28 | 0.034 | 0.39 (0.03 to 0.66) |
| LPC(22:4) | -2.08 | 28 | 0.047 | -0.37 (-0.006 to -0.64) |
| **Th17 %** |  |  |  |  |
| LPEI(16:0) | -2.52 | 26 | 0.018 | -0.44 (-0.08 to -0.70) |
| DG(18:0/18:1) | -2.11 | 26 | 0.045 | -0.38 (-0.01 to -0.66) |
| PI(16:1/18:1) | -2.09 | 26 | 0.046 | -0.38 (-0.01 to -0.66) |
| **Effector CD4)^+^ T cell %** |  |  |  |  |
| PI(16:0/18:1) | -2.11 | 27 | 0.044 | -0.38 (-0.01 to -0.65) |
| **Th1 %** |  |  |  |  |
| PI(16:0/20:4) | 2.76 | 26 | 0.011 | 0.48 (0.12 to 0.72) |
| PI(18:1/20:4) | 2.74 | 26 | 0.011 | 0.47 (0.12 to 0.72) |
| LPE(16:0) | 2.55 | 26 | 0.017 | 0.45 (0.09 to 0.70) |
| LPE(18:0) | 2.36 | 26 | 0.026 | 0.42 (0.05 to 0.69) |
| PI(16:1/18:1) | 2.32 | 26 | 0.028 | 0.41 (0.05 to 0.68) |
| **MSIS physical** |  |  |  |  |
| LPC(22:4) | 2.66 | 27 | 0.013 | 0.46 (0.11 to 0.70) |
| LPC(18:2) | 2.41 | 27 | 0.023 | 0.42 (0.06 to 0.68) |
| LPC(20:4) | 2.40 | 27 | 0.024 | 0.42 (0.06 to 0.68) |
| PI(16:0/20:4) | 2.30 | 27 | 0.030 | 0.40 (0.04 to 0.67) |
| LPC(16:0) | 2.23 | 27 | 0.034 | 0.39 (0.03 to 0.66) |
| LPE(18:1) | 2.14 | 27 | 0.042 | 0.38 (0.02 to 0.66) |
| LacCer(d18:1/22:1) | -2.13 | 27 | 0.043 | -0.38 (-0.01 to -0.65) |
| **MFIS cognitive** |  |  |  |  |
| LacCer(d18:1/22:1) | -2.28 | 27 | 0.031 | -0.40 (-0.04 to -0.67) |
| **MFIS psychosocial** |  |  |  |  |
| LPE(18:1 (*FDR q=0.032) | 3.41 | 27 | 0.002 | 0.55 (0.23 to 0.76) |
| LPC(18:2) | 2.46 | 27 | 0.021 | 0.43 (0.07 to 0.69) |
| PI(16:0/18:1) | 2.35 | 27 | 0.026 | 0.41 (0.05 to 0.68) |
| LPC(22:4) | 2.28 | 27 | 0.031 | 0.40 (0.04 to 0.67) |

|  | **S-statistic** |  | **P-value** | **Spearman's correlation coefficient** |
| --- | --- | --- | --- | --- |
| **Plasma B cell number** |  |  |  |  |
| LPC(18:2) | 5686 |  | 0.032 | -0.40 |
| **PASAT 3 sec average** |  |  |  |  |
| LPE(16:0) | 1977 |  | 0.004 | 0.51 |
| LPE(18:0) | 2447 |  | 0.033 | 0.40 |
| LPC(18:1) | 2471 |  | 0.036 | 0.39 |
| PI(16:0/18:1) | 2538 |  | 0.045 | 0.37 |
| LPC(16:0) | 2567 |  | 0.050 | 0.37 |
