## Supplementary material for "RANDOMIZED CONTROLLED TRIAL OF INTERMITTENT CALORIE RESTRICTION IN PEOPLE WITH MULTIPLE SCLEROSIS": Supp. Table 9

|  |  | **iCR** | | **Control** | |  | **iCR vs Control** | |
| --- | --- | --- | --- | --- | --- | --- | --- | --- |
|  |  | **Mean change (95% CI)** | ***P*-value** | **Mean change**  **( 95% CI)** | ***P*-value** |  | **Mean difference**  **( 95% CI)** | ***P-*value** |
| **CLINICAL OUTCOME MEASURES** | | | | | | | | |
| **Expanded disability status scale (EDSS)** | ***0-6 weeks*** | 0.328  (-0.012, 0.668) | ***0.04*** | -0.487  (-0.837, -0.137) | ***0.01*** | ***Week***  ***6*** | -0.761  (-1.326, -0.195) | ***0.01*** |
|  | ***0-12 weeks*** | 0.097  (-0.251, 0.444) | 0.41 | -0.277  (-0.626, 0.073) | 0.13 | ***Week***  ***12*** | -0.319  (-0.889, 0.251) | 0.22 |
| **Symbol Digit Modalities test (SDMT- average)** | ***0-6 weeks*** | -3.512  (-6.388, -0.636) | ***0.02*** | -2.622  (-5.513, 0.268) | 0.09 | ***Week***  ***6*** | 0.597  (-5.746, 6.941) | 0.23 |
|  | ***0-12 weeks*** | -6.277  (-9.153, -3.401) | ***< 0.0001*** | -1.419  (-4.31, 1.472) | 0.40 | ***Week***  ***12*** | 4.566  (-1.778, 10.909) | ***0.03*** |
| **Timed 25-foot walk (average)** | ***0-6 weeks*** | -0.009  (-0.218, 0.201) | 0.84 | 0.184  (-0.031, 0.398) | 0.09 | ***Week***  ***6*** | 0.367  (-0.384, 1.118) | 0.85 |
|  | ***0-12 weeks*** | 0.086  (-0.124, 0.295) | 0.49 | -0.075  (-0.29, 0.14) | 0.50 | ***Week***  ***12*** | 0.014  (-0.737, 0.765) | 0.71 |
| **9-Hole Peg Test (average)** | ***0-6 weeks*** | 0.586  (-0.278, 1.45) | 0.21 | 0.603  (-0.283, 1.489) | 0.18 | ***Week***  ***6*** | -0.168  (-2.580, 2.244) | 0.57 |
|  | ***0-12 weeks*** | 1.055  (0.191, 1.919) | ***0.02*** | 1.071  (0.185, 1.956) | ***0.02*** | ***Week***  ***12*** | -0.169  (-2.582, 2.243) | 0.57 |
| **Paced Auditory Serial Addition Test (PASAT-3-seconds)** | ***0-6 weeks*** | -3.465  (-5.9, -1.029) | ***0.007*** | -3.597  (-6.175, -1.02) | ***0.007*** | ***Week***  ***6*** | 1.429  (-5.211, 8.070) | 0.45 |
|  | ***0-12 weeks*** | -4.759  (-7.194, -2.323) | ***< 0.001*** | -3.724 (-6.302, -1.147) | ***0.005*** | ***Week***  ***12*** | 2.597  (-4.044, 9.237) | 0.28 |
| **MS Functional Composite (z-score)** | ***0-6 weeks*** | -0.149  (-0.247, -0.05) | ***0.005*** | -0.15  (-0.251, -0.049) | ***0.006*** | ***Week***  ***6*** | 0.071  (-0.224, 0.366) | 0.63 |
|  | ***0-12 weeks*** | -0.208  (-0.307, -0.109) | ***< 0.001*** | -0.179  (-0.281, -0.078) | ***0.001*** | ***Week***  ***12*** | 0.101  (-0.195, 0.396) | 0.52 |
| **PATIENT-REPORTED OUTCOMES** | | | | | | | | |
| **Modified Fatigue Impact Scale (MFIS)** |  |  |  |  |  |  |  |  |
| **Physical subscale** | ***0-6 weeks*** | 0.483  (-0.084, 1.05) | 0.09 | -0.163  (-0.733, 0.408) | 0.42 | ***Week***  ***6*** | 0.218  (-0.772, 1.208) | 0.96 |
|  | ***0-12 weeks*** | 0.517  (-0.063, 1.096) | 0.07 | -0.163  (-0.733, 0.408) | 0.42 | ***Week***  ***12*** | 0.184  (-0.813, 1.182) | 0.99 |
| **Cognitive subscale** | ***0-6 weeks*** | 1.52  (-0.609, 3.648) | 0.15 | 1.188  (-0.947, 3.323) | 0.33 | ***Week***  ***6*** | 2.878  (-2.223, 7.979) | 0.30 |
|  | ***0-12 weeks*** | 2.548  (0.371, 4.726) | ***0.02*** | 1.306  (-0.829, 3.441) | 0.28 | ***Week***  ***12*** | 1.967  (-3.155, 7.088) | 0.41 |
| **Psychological subscale** | ***0-6 weeks*** | 2.088  (0.126, 4.05) | ***0.04*** | 0.918  (-1.049, 2.885) | 0.43 | ***Week***  ***6*** | 0.894  (-4.422, 6.209 | 0.25 |
|  | ***0-12 weeks*** | 2.839  (0.832, 4.847) | ***0.006*** | 0.977  (-0.99, 2.944) | 0.40 | ***Week***  ***12*** | 0.201  (-5.131, 5.534) | 0.32 |
| **MSIS (mental)** | ***0-6 weeks*** | 4.297  (0.638, 7.957) | ***0.02*** | 2.143  (-1.448, 5.734) | 0.29 | ***Week***  ***6*** | 1.456  (-7.912, 10.824) | 0.24 |
|  | ***0-12 weeks*** | 7.357  (3.613, 11.102) | ***< 0.001*** | -0.645  (-4.409, 3.118) | 0.65 | ***Week***  ***12*** | -4.392  (-13.860, 5.075) | 0.70 |
| **MSIS (physical)** | ***0-6 weeks*** | 2.765  (0.407, 5.122) | ***0.02*** | 1.906  (-0.455, 4.266) | 0.14 | ***Week***  ***6*** | 0.058  (-7.982, 8.097) | 0.69 |
|  | ***0-12 weeks*** | 1.02  (-1.392, 3.431) | 0.36 | 1.000  (-1.361, 3.36) | 0.47 | ***Week***  ***12*** | 0.897  (-7.159, 8.952) | 0.44 |

Mean changes within group and mean difference between groups are calculated based on the raw means. Changes over time within group were calculated subtracting the values at week 6 or 12 to the values at baseline. Therefore, negative values correspond to an increase. Mean differences were calculated as follows: iCR mean at 6 (or 12) weeks minus control mean at 6 (or 12) weeks. P values are adjusted for age, sex and MS disease modifying therapy use. Mean reductions or increases reported in the table are not adjusted. MSIS: Multiple Sclerosis Impact Scale.
